# Projections of the impact of transmission-reducing pediatric influenza vaccine in South Africa using high resolution immunologic and infection data

**DOI:** 10.1101/2025.04.04.25325228

**Authors:** Jackie Kleynhans, Cécile Viboud, Molly Sauter, Stefano Tempia, Nicole Wolter, Jocelyn Moyes, Anne von Gottberg, Lorens Maake, Alexandra Moerdyk, Cheryl Cohen, Kaiyuan Sun

## Abstract

**Background:** Traditional influenza vaccination programs focus on preventing severe illness through the vaccine’s direct effect. However, vaccinating children may substantially reduce overall burden by lowering transmission, especially in high infection settings and when effective transmission reducing vaccines become available.

**Methods:** To evaluate the impact of a transmission-reducing vaccination campaign, we developed a Susceptible-Latent-Infectious-Recovered (SLIR) compartmental transmission model calibrated with high-resolution virological and immunological data sourced from a multiyear household cohort study in urban and rural South Africa. The mode explicitly accounted for variations in susceptibility, infectiousness, and viral shedding duration by age group, pre-season hemagglutination inhibition antibody titer and influenza subtype/lineage. Using this framework, we simulated pediatric vaccination scenarios targeting children aged 6 months – 5 years or 6-12 years, varying both vaccine coverage and the vaccine’s effectiveness in reducing susceptibility and transmission. Scenarios were compared to no-vaccination counterfactuals to project reductions in infections, illness episodes, hospitalizations, and deaths in South Africa.

**Results:** Our model projects that vaccinating 50% of children aged 6 months - 5 years or 6-12 years with a vaccine that is 50% effective in reducing both susceptibility and viral shedding could lower the overall infection incidence by 58-60% and 48%-51%, respectively. Illness episodes would be reduced by 20-60%, hospitalizations by 26-64%, and deaths by 21-59% across all age groups. The target age group among children that resulted in the highest population-wide vaccination benefits varied with the circulating lineage, season, and urbanization level.

**Conclusions:** Our work demonstrates that pediatric immunization with transmission-reducing vaccines holds significant potential for lowering community influenza burden, particularly in high transmission settings. Our modeling framework offers a practical translational tool for anticipating the impact of new vaccines in low and middle-income settings.

## Introduction

Influenza remains a significant cause of morbidity and mortality globally despite well-established influenza vaccine programs. Influenza-associated lower respiratory tract infections are globally responsible for 124 hospitalizations and 1.9 deaths per 100,000 population annually, with higher rates (156 hospitalizations and 2.7 deaths per 100,000 population) in sub-Saharan Africa [1]. In South Africa, where the influenza burden pyramid has been well-characterized, an estimated 10.7 million episodes of influenza-associated illness occur annually, 1.2% of these are severe and 0.1% result in death [2], amounting to an annual cost of US$270.5 million [3]. While children younger than 13 years in this setting have very high attack rates (51-66 per 100 person seasons) [4], severe illness and mortality is concentrated at the extremes of age, and among people living with HIV and other underlying conditions [5].

Seasonal influenza vaccination reduces the risk of severe influenza-associated illness and death in vaccinated individuals [6]. Globally, vaccination programs have focused on high risk groups, including pregnant woman, children aged 6-59 months, the elderly and those with comorbidities [7]. Although forty-five percent of South Africa’s population falls into high-risk groups targeted for influenza vaccination [8], the public national influenza program procures only a limited number of vaccines, prioritizing individuals in these groups and healthcare workers. Yet, seasonal influenza coverage in South Africa remains low, estimated at 4.6% of this target population in 2018 [9].

An alternative influenza control strategy could be considered, which would harness the transmission-reducing effects of vaccines. By prioritizing vaccination in age groups with high attack rates and significant roles in viral transmission, this approach would aim to reduce influenza circulation at the community level. High-risk individuals who remain unvaccinated benefit indirectly from this policy through reduced exposure to influenza – the so-called indirect effect. To date, however, this transmission-focused strategy has been primarily explored through mathematical modeling [10–13], with real-world assessments mostly focused in high income countries such as Canada, Japan, Russia, the United States of America and the United Kingdom [14–20]. While a school-based influenza vaccination program has been implemented in England [19, 20], this approach has not yet been implemented in any low and middle-income countries, where influenza burden is the highest [1], and potential indirect impacts may be greater.

Two major challenges impede the practical implementation of influenza immunization programs designed to harness the vaccine’s indirect protective effects. First, current vaccines are primarily designed to prevent severe outcomes, and their effects on transmission are rarely assessed in clinical trials. Second, vaccination strategies relying on indirect vaccine effects should target the population groups that contribute the most to influenza transmission, regardless of clinical severity. Although children have been identified as major transmitters by a number of symptom-based surveillance and epidemiological studies [4, 21, 22], these studies often overlook asymptomatic and subclinical infections that also drive transmission. As a result, the potential benefits of pediatric vaccination may be under- or overestimated depending on the age group in which much asymptomatic infections occur, and the findings may not generalize to regions with different population demographics and influenza dynamics.

In this study, we introduce a comprehensive modeling framework to assess the potential impact of transmission-reducing vaccines targeting children. By leveraging detailed infection, serological and contact data from South African household cohorts, our model integrates the roles of asymptomatic transmission, variations in susceptibility and infectiousness by age and pre-existing HAI titers, and mixing patterns [4, 23, 24]. By systematically simulating a range of vaccine efficacy profiles and immunization coverages against different influenza subtypes/strains, we identify strategies that could markedly reduce the overall burden of influenza by targeting the high-transmitting pediatric population. Our approach not only addresses critical gaps in the current assessments of vaccine impact on transmission but also provides actionable tools for optimizing vaccination strategies.

## Materials and Methods

To evaluate the potential impact of transmission-reducing influenza vaccines, we developed a transmission and vaccine-impact modeling framework calibrated to detailed infection and serological data from longitudinal household cohorts in South Africa. First, we describe the underlying cohort data. Next, we introduce the structure, parameterization and calibration of our transmission model, along with assumptions underpinning our vaccine intervention simulations. Finally, we outline our methodology to project the amount of disease burden averted by implementing a pediatric vaccination program in South Africa.

### Household cohort study for model calibration

We used data from a prospective household cohort study where 50 households were randomly selected annually from each of an urban and a rural community and intensively followed up for six to ten months spanning the 2016-2018 influenza seasons in South Africa (PHIRST study [4]). Briefly, participants were visited twice a week for the collection of nasopharyngeal specimens for PCR testing of influenza viruses A and B irrespective of symptoms, with typing to A(H1N1)pdm09, A(H3N2), B/Victoria and B/Yamagata for all positive samples. In addition, pre-season serum was collected to test for presence of influenza antibodies for each subtype/lineage, using hemagglutination inhibition (HAI) assays with viruses that circulated during the period (A/South Africa/2517/2016 A(H1N1)pdm09, A/Singapore/INFIMH-16-0019/2016 A(H3N2), B/South Africa/R3037/2016 (B/Victoria), and B/South Africa/R05631/2017 (B/Yamagata)). In 2018, a contact survey was performed to collect age-based contact rates in the cohort [25].

### Model

We constructed a deterministic Susceptible-Latent-Infectious-Recovered (SLIR) transmission model, where individuals move between susceptible (S), latent (L), infectious (I) and recovered (R) compartments. The model allowed for both susceptibility to infection and infectiousness to vary by age and influenza antibody titers (hereafter referred to as titers), with age- and titer-specific susceptibility and infectiousness calibrated on our prior analysis of shedding dynamics and infection risk in the South African household cohort [24]. The compartmental model was further stratified by age (<6 months, 6 months – 5 years, 6-12 years, 13-18 years, 19-44 years, 45-64 years and ≥65 years), and pre-season titer for each subtype/lineage (<1:40, 1:40-1:80, ≥1:160).

The impact of age and antibody titers on transmissibility was modeled by assigning different latent periods, infectious periods, and relative infectiousness depending on age and titer, following prior analysis ([24] and Supplementary Excel File 1). Within each age and titer group, the average latent period was defined as the sum of half the duration of the viral shedding proliferation phase and the interval from infection to the first detectable PCR positivity [26]; the average infectious period was half of the shedding duration; and the relative infectiousness was approximated by the ratios of peak viral loads (Fig. S1). Compared to the original estimates in Sauter *et al.* [24], we updated the age categories and discretized titer as a categorical variable to accommodate the parameterization of the SLIR model used in this paper. Adjusted susceptibility estimates are provided in Supplementary Excel File 1.

Separate models were built for each cohort study site (rural: Agincourt, urban: Klerksdorp), season (2016, 2017, 2018) and subtype/lineage (A(H1N1)pdm09, A(H3N2), B/Victoria and B/Yamagata). In some of our analyses, we projected influenza epidemics separately for each subtype/lineage, and in others we combined projections for all subtypes/lineages co-circulating the same year to reflect the “total” annual influenza burden (by summing infections).

Population size (*N*) was set at 10,000,000, with the age distribution reflecting the age distribution of the PHIRST cohort for the respective sites (Figures 1A). The site-specific mixing patterns (Figures 1B) among different age groups were based on a cross-sectional contact survey done in the 2018 cohort [25]. Within each age group, the proportion of individuals with each antibody titer (*h*) was based on the cohort site- and year-specific pre-season titers for the respective subtype/lineage (Figures 1C).

**Fig. 1.**
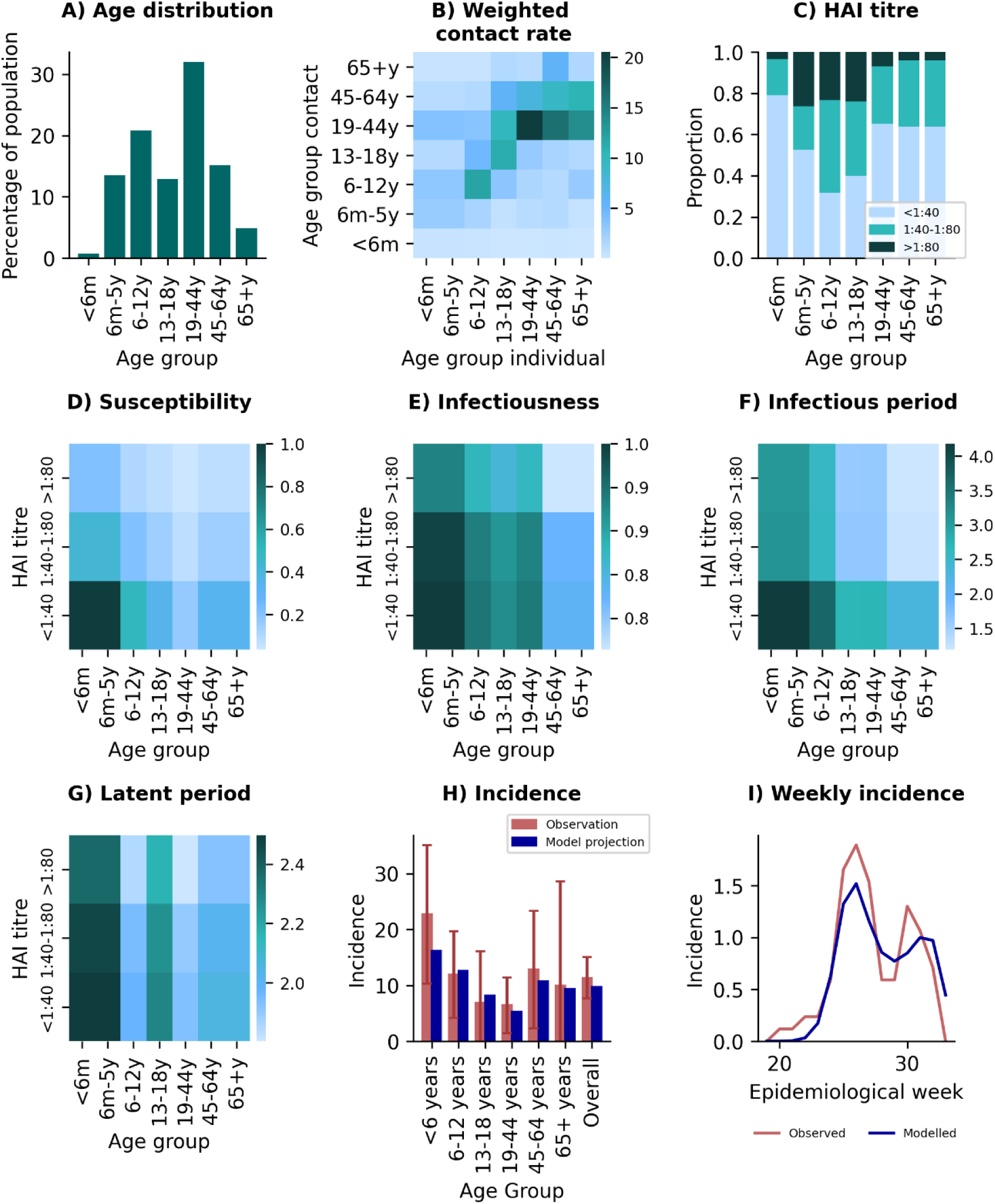
Model inputs (A-G) and calibration targets (H-I) for A(H1N1)pdm09, urban site, 2018. A) Model population age structure (m – months, y - years). B) Cumulative contact rates between age groups at home, school, work and within the community, weighted by calibrated omega (matrix weight). C) Proportion of population with pre-season hemagglutination inhibition assay (HAI) titers for A(H1N1)pdm09 within each age group. D) Susceptibility (α, risk of infection), E) infectiousness (β, minimum Ct value as proxy for viral load), F) infectious period (µ, ½ duration of shedding) and G) latent period (σ, infection to infectiousness) by age and pre-season HAI titers. H) Observed and modelled incidence per 100 population (with 95% confidence intervals) by age group and I) weekly observed and modelled A(H1N1)pdm09 incidence used for model calibration.

We did not consider antibody (immunity) waning during the season, or cross-protection of antibodies across subtypes/lineages.

To project the potential impact of vaccination, the model was further broken down by vaccination status (vaccinated or not at the start of the season), in addition to age and antibody titers. This allowed us to model the potential impact of vaccination on susceptibility against infection as well as shedding kinetics and infectiousness.

The equations of this SLIR model were:

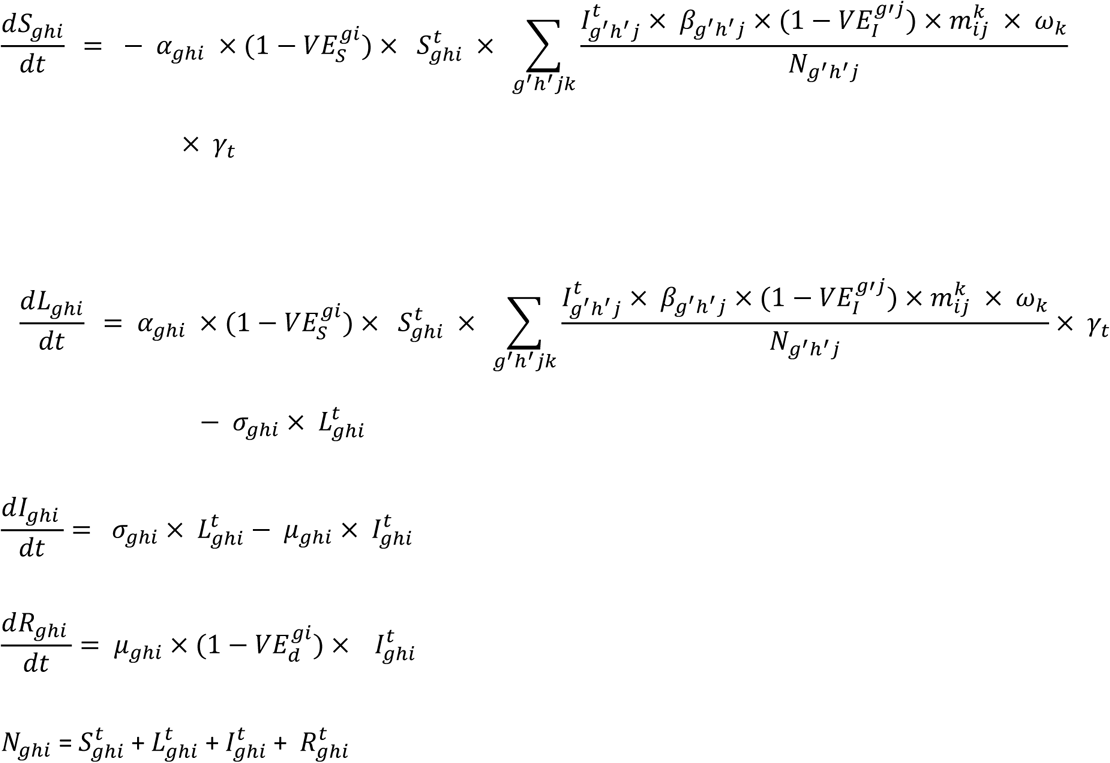

Where 𝑁_𝑔ℎ𝑖_ is the number of individuals with vaccination status *g*, pre-season titer *h* in age group 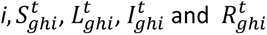 represent the number of individuals in the Susceptible, Latent, Infectious and Recovered compartments at time point t.

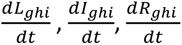 are the rate at which individuals move between compartments. 𝛼_𝑔ℎ𝑗_ is the susceptibility, 𝛽_𝑔ℎ𝑗_ is the infectiousness, 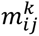 is the daily contact rate between individuals in age group *i* with age group *j* in contact setting *k (e.g.* household, school, workplace, or community). 𝜔_𝑘_ is the relative weight of contacts in setting *k*, as per-contact transmission risk is setting dependent. 𝛾_𝑡_ is the baseline transmissibility at time point *t*. 𝜎_𝑔ℎ𝑖_ is the 1/latency period, 𝜇_𝑔ℎ𝑖_ the recovery rate (1/duration of shedding). All individuals in 𝑅 compartment behave the same, irrespective of their vaccine, pre-season titer level, and age, with no transition out of 𝑅 compartment.

𝑉𝐸_𝑠_ and 𝑉𝐸_𝑖_ represent the vaccine effectiveness on susceptibility and infectiousness (viral load), respectively. 𝑉𝐸_𝑖_ and 𝑉𝐸_𝑑_ collectively is the vaccine effectiveness against transmission. In the current models, 𝑉𝐸_𝑖_ was set to 0 and 𝑉𝐸_𝑑_ is considered the VE against transmission, although the framework would allow 𝑉𝐸_𝑖_ to be adjusted.

To initialize the model, we set the initial number of individuals infected 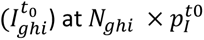, where 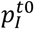 is the proportion of individuals initially infected (the same across stratification) at the beginning of each influenza season, which is a free parameter to be estimated during model calibration. The initial number of susceptible 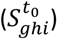 was 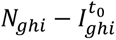, and the initial number in the latent 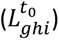 and recovered 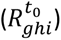 compartments were 0. 𝑡_0_ represents the starting time of the epidemic season.

A description of model parameters is provided in Table S1 and actual parameter values used for individual models are provided in Supplementary Excel file 1. Calibrated model parameters included the initial proportion infected 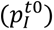, the time-varying transmissibility (𝛾_𝑡_, unique to each strain and year), and a contact matrix weight (𝜔, shared across strains and years). All of the model parameters were based on estimates directly derived from the PHIRST cohort, as previously described in Sauter *et al.* [24] and Kleynhans *et al.* [25].

### Model calibration

Models were calibrated to match site-, season-, and subtype-specific infection data from PHIRST based on PCR-confirmed influenza. We calibrated the model against 1) weekly all-age incidence of infections (where infection was assessed by twice weekly reverse transcription real-time polymerase chain reaction (rRT-PCR) tests on nasopharyngeal swabs irrespective of symptoms [4]) and 2) age-specific cumulative seasonal infection attack rate (Table S2).

For each subtype/lineage model for each site and year, we calculated the squared residual of the weekly incidence (𝑅𝑒𝑠𝑖𝑑𝑢𝑎𝑙^2^_𝑤𝑘_𝑖𝑛𝑐_) and age-based and overall attack rates (𝑅𝑒𝑠𝑖𝑑𝑢𝑎𝑙^2^_𝑎𝑔𝑒_𝑜𝑣𝑒𝑟𝑎𝑙𝑙_𝑖𝑛𝑐_) between the model and observed (PHIRST) data. Model error 𝑅𝑒𝑠𝑖𝑑𝑢𝑎𝑙^2^_subtype/lineage_ was calculated as the product of 𝑅𝑒𝑠𝑖𝑑𝑢𝑎𝑙^2^_𝑤𝑘_𝑖𝑛𝑐_ and 𝑅𝑒𝑠𝑖𝑑𝑢𝑎𝑙^2^_𝑎𝑔𝑒_𝑜𝑣𝑒𝑟𝑎𝑙𝑙_𝑖𝑛𝑐_. We then calculated an overall observational error as the product of 𝑅𝑒𝑠𝑖𝑑𝑢𝑎𝑙^2^_subtype/lineage_ across all lineages and years, which we minimized. We only modelled and considered subtypes/lineages with an attack rate above 2/100 population per season in the combined models (Table S2). For calibration purposes, we set vaccine coverage at 0 for all ages since vaccination was not routinely used in the cohort study population (<1% [4]).

### Modeling vaccine impact on infections

To assess the potential impact of different coverage levels, age groups targeted, and vaccine profiles, we ran year-, site- and subtype/lineage-specific models for different vaccination scenarios, exploring different levels of vaccine effectiveness for 𝑉𝐸_𝑠_ and 𝑉𝐸_𝑑_ and vaccine coverage in 6 months – 5 years, 6-12 years or combined 6 months – 12 years. The calibrated model, with vaccine coverage set to 0, represented the counterfactual no-vaccination base model.

Vaccine impact was calculated as the absolute (𝐼𝑛𝑐𝑖𝑑𝑒𝑛𝑐𝑒_𝑏𝑎𝑠𝑒 𝑚𝑜𝑑𝑒𝑙_ − 𝐼𝑛𝑐𝑖𝑑𝑒𝑛𝑐𝑒_𝑉𝑎𝑐𝑐𝑖𝑛𝑒 𝑚𝑜𝑑𝑒𝑙_) and percentage reduction in incidence 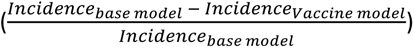 by age group and overall between the model with (vaccine model) and without (base model) vaccination. We also calculated infections averted per dose as 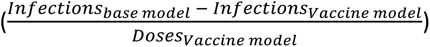

To calculate the overall influenza incidence and vaccine impact, we created ‘typical’ influenza seasons, through combining the infections from the respective subtypes/lineages reported to co-circulate in a given site and year and divided by the population size. These seasonal incidence estimates were used in the vaccine impact calculations.

### Projections of vaccine reductions in disease burden

To project the overall impact of different vaccination strategies on influenza burden, we used age-based multipliers developed for the South African population to derive the number of (symptomatic) illness episodes, hospitalizations and deaths based on projected infections by our model (Table S3). To estimate the number of symptomatic illness episodes, we used the symptomatic fraction reported in the PHIRST cohort [4]. To estimate hospitalizations and deaths, we used the proportion of infections resulting in hospitalizations or deaths from a disease burden study in South Africa [2]. We adapted the age breakdowns of the disease burden study [2] to our transmission model (Table S3).

To account for inter-annual fluctuations in influenza disease burden, we averaged illness outcomes over the three study years to obtain annual rates. This was done for the counterfactual and for each vaccine coverage scenario, with a VE set at 50% against both susceptibility and duration of shedding for illustration. We assumed that the vaccine had no additional impact on illness severity beyond the effect on preventing infection and reducing transmission. In other words, we ignored the potential effect of the vaccine on reducing the risk of developing severe illness among breakthrough infections. The percentage reduction of each vaccine scenario and illness outcome was calculated as 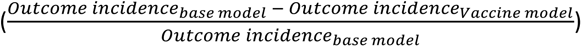, with illness outcome representing symptomatic illness, hospitalizations or deaths.

Model simulation and analyses were performed using Python v3.9.12.

## Results

In Fig. 1 we summarize the model inputs and calibration targets for the model. The age distribution in the rural and urban sites differed in the South African household cohorts, with a higher proportion of children aged 6 months – 18 years at the rural site (Fig. S2 A-B). These differences likely drove the differences in contact patterns between the two sites, with much higher contact rates in the rural site among children and teenagers (Fig. S2 C-J). Pre-season antibody titers differed across age groups, years and subtypes/lineages (Fig. S3-4). Susceptibility (Fig. 1D, S2), infectiousness (Fig. 1E, S6), infectious period (Fig. 1F, S7) and latent period (Fig. 1G, S8) estimates followed a decreasing gradient with the increase in age and titer. These factors likely contributed to the year-to-year and site variations in subtypes/lineages circulating, with differences in incidence rates observed by season and site (Fig. 1H-I, S9-10). Our models performed well to estimate age-based incidence within the 95% confidence intervals of observed data, and to track weekly epidemic curves each season (Fig. 1H-I, S9-10).

In Fig. 2, we illustrate the impact of vaccine modeling results for 2018 at the urban site with 50% of children aged 6-12 years vaccinated, with additional results for other vaccine scenarios, sites and years shown in Fig. S9-26. Our model predicted that reductions in influenza incidence varied according to vaccine impact on susceptibility and viral shedding, with both mechanisms contributing comparably (Fig. 2A). For an exemplar season marked by co-circulation of A(H1N1)pdm09 and B/Victoria (as in 2018), we observed a 42% reduction in influenza incidence in all ages with a VE of 50% against duration of shedding and no effect on susceptibility, compared to a 53% reduction with 50% VE against susceptibility and no effect on shedding. Assuming a 50% VE against both duration of shedding and susceptibility resulted in a 67% reduction in influenza incidence. We used this VE assumption for the remaining analyses in the paper.

**Fig. 2.**
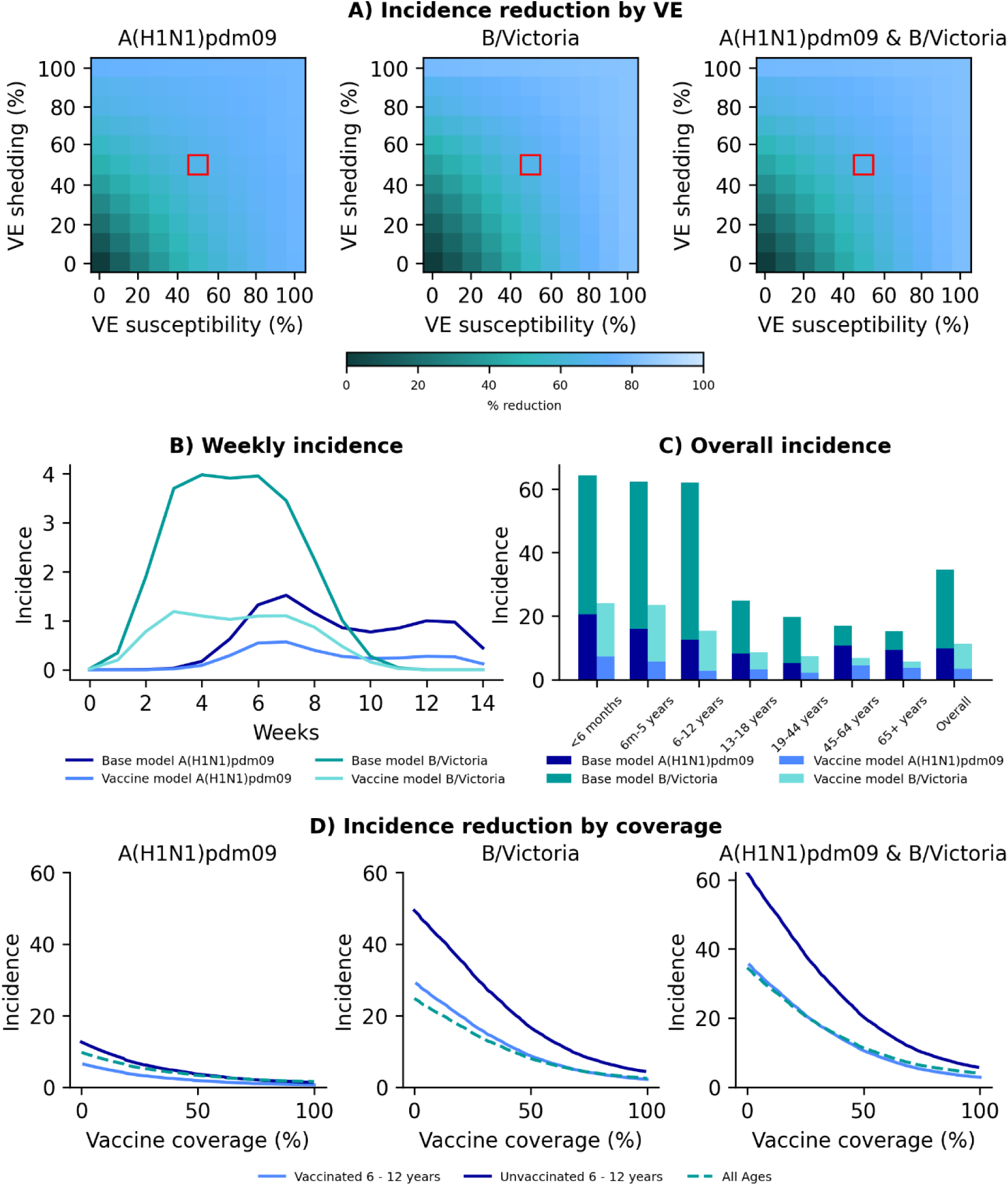
Vaccine model results for vaccinating children aged 6-12 years, urban site, 2018 season. A) Percentage reduction in incidence at differing levels of vaccine effectiveness (VE) against susceptibility (x axis) and duration of shedding (y axis) when vaccinating 50% of the target age group. Red block denotes VE chosen (50%) for subsequent analysis. B) Modelled weekly incidence per 100 population of individual influenza subtype/lineage in base model (no vaccination) and vaccine model. C) Overall incidence of influenza per 100 population subtype/lineage by age in base and vaccine model. Both B and C based on 50% coverage in target age group with vaccine with VE of 50% against susceptibility and duration of shedding. D) Incidence of influenza per 100 population subtype/lineage (left and middle) and combined season (right) at varying vaccine coverage levels in the target age group with vaccine with VE of 50% against susceptibility and duration of shedding.

The weekly incidence in vaccination scenarios initially followed the same trajectory as the no-vaccination scenario, diverging primarily at the peak—where the magnitude was reduced, though its timing remained consistent (Fig. 2B). The overall percentage reduction depended on age groups targeted for vaccination, with greatest benefits observed when the age group with highest incidence was immunized. Notably, the age group with highest incidence varied based on the circulating subtypes/lineages, year and site. For example, during the 2018 urban season, children aged 6-12 years experienced an attack rate of 62 per 100 persons, and vaccinating this age group reduced the overall incidence by 67% (Fig. 2C). In contrast, the urban 2017 season, characterized by circulation of A(H3N2) and B/Yamagata, recorded extremely high attack rates in children aged 13-18 years (98 per 100 persons, which includes multiple infections), 2 to 3 times higher than in children aged 6 months - 12 years or 6-12 years. In this season, we find more modest incidence reduction in all ages when vaccinating children aged 6 months – 5 years or 6 – 12 years, at 9% and 34% respectively (Fig. S14 and S15). As expected, higher vaccine coverage led to greater reductions in incidence, with a rapid decline in incidence as coverage increased up to 50%, followed by more gradual benefits beyond that level (Fig. 2D). Additional model results for all seasons, vaccination strategies and both sites are presented in S11-S27.

To better understand the potential range of vaccine benefits, we summarized infection reductions by targeted age group, year and site (Fig. 3). We assumed 50% vaccine coverage using a vaccine with 50% VE in reducing both susceptibility to infection and the duration of viral shedding. Projected vaccine benefits varied substantially by year and study site, ranging from a 9% reduction in incidence in all ages when vaccinating children aged 6 months – 5 years in the urban site in 2017, to a 93% reduction when vaccinating children aged 6 months – 12 years in the 2018 season in the rural site (Fig. 3). This correspond to a range of 0.5 to 2 infections averted for every dose administered. When only one of the two age groups was targeted, we saw the greatest percentage reduction when vaccinating children aged 6-12 years in the rural site in the 2018 season when A(H1N1)pdm09 and B/Victoria were circulating, with a 69% reduction in incidence and 2.5 infections averted per dose administered (Fig. 3B). In both sites, we observed the lowest reductions in incidence when A(H3N2) and B/Yamagata were circulating, and highest when A(H1N1)pdm09 and B/Victoria were circulating. There were also differences in the level of reduction between the two sites even when the same subtypes/lineages were circulating (Fig. 3A, B). For example, in 2018, although both sites experienced A(H1N1)pdm09 and B/Victoria, the urban site benefited more from vaccinating children aged 6–12 years, whereas the rural site saw greater effects from vaccinating children aged 6 months – 5 years.

**Fig. 3.**
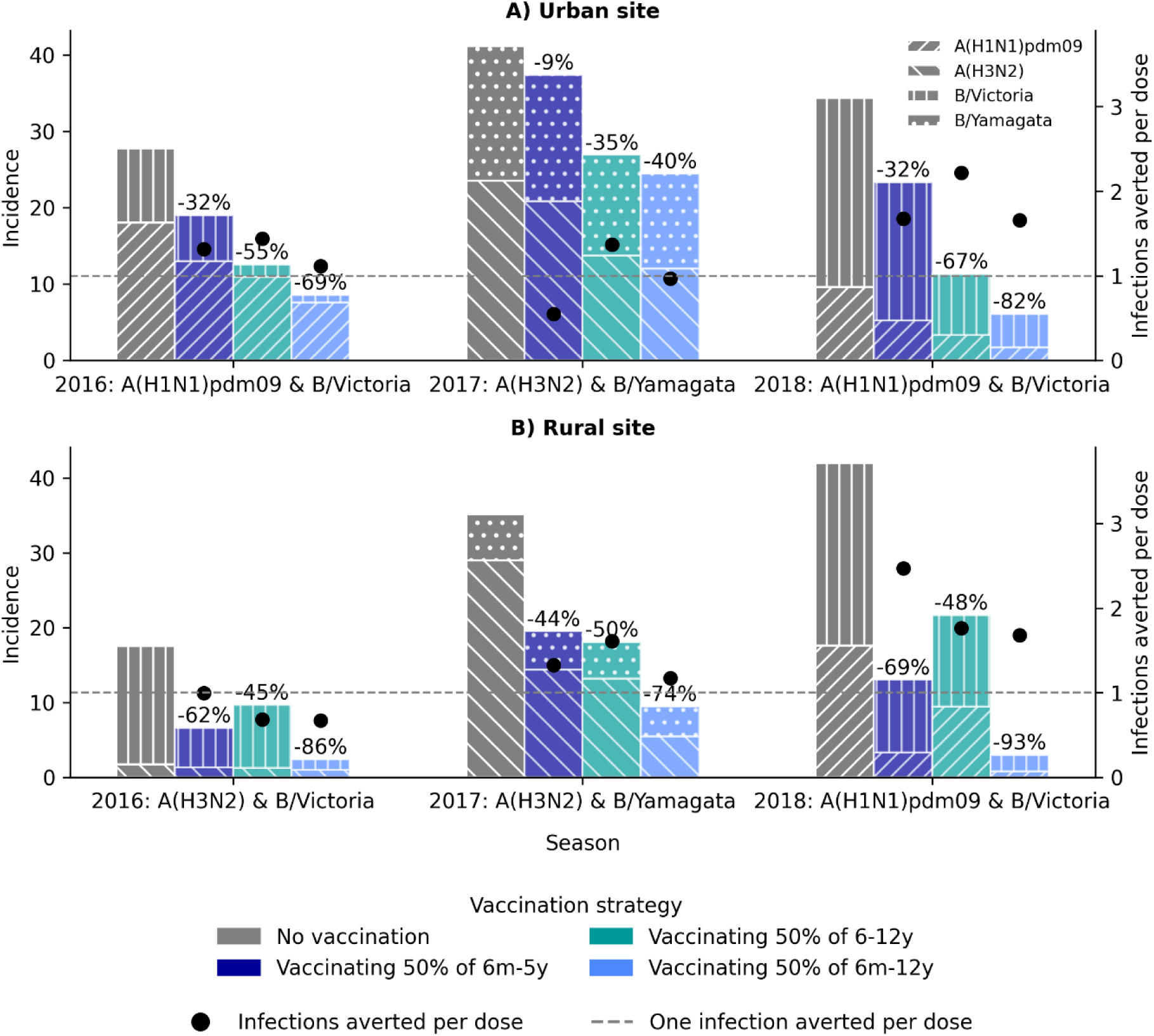
Modelled influenza incidence per 100 population (bars, primary axis) in the base model (no vaccination) and three scenarios of vaccinating 50% of the target age group (6 months to 5 years, 6-12 years and 6 months – 12 years) with a vaccine with 50% effectiveness against susceptibility and duration of shedding; infections averted per dose (dots, secondary axis); reference for one infection averted per dose (grey dotted line, the secondary axis); percentage reduction (text at edge of bar), at A) urban and B) rural site, 2016-2018 seasons, South Africa

Given the heterogeneity of influenza circulation and vaccine effects across different seasons, we summarized the putative benefits of a vaccination program on infections, symptomatic illness, hospitalizations and deaths by averaging results over 3 seasons (Fig. 4). Across all illness outcomes and for both metrics of percentage reduction and burden averted per dose administered, the strategy to vaccinate children aged 6 months – 5 years fared better in the rural site, while the strategy to vaccinate children aged 6-12 years fared better in the urban site (Fig. 4B). Depending on the outcome considered, the influenza disease burden could be reduced by 58-64% in the rural site, and 43-51% in the urban site. For both sites, irrespective of strategy, at least 1.2 and 0.6 infections and illness episodes could be averted per dose, corresponding to at least 8 hospitalizations and 0.4 deaths averted per 1000 doses administered.

**Fig. 4.**
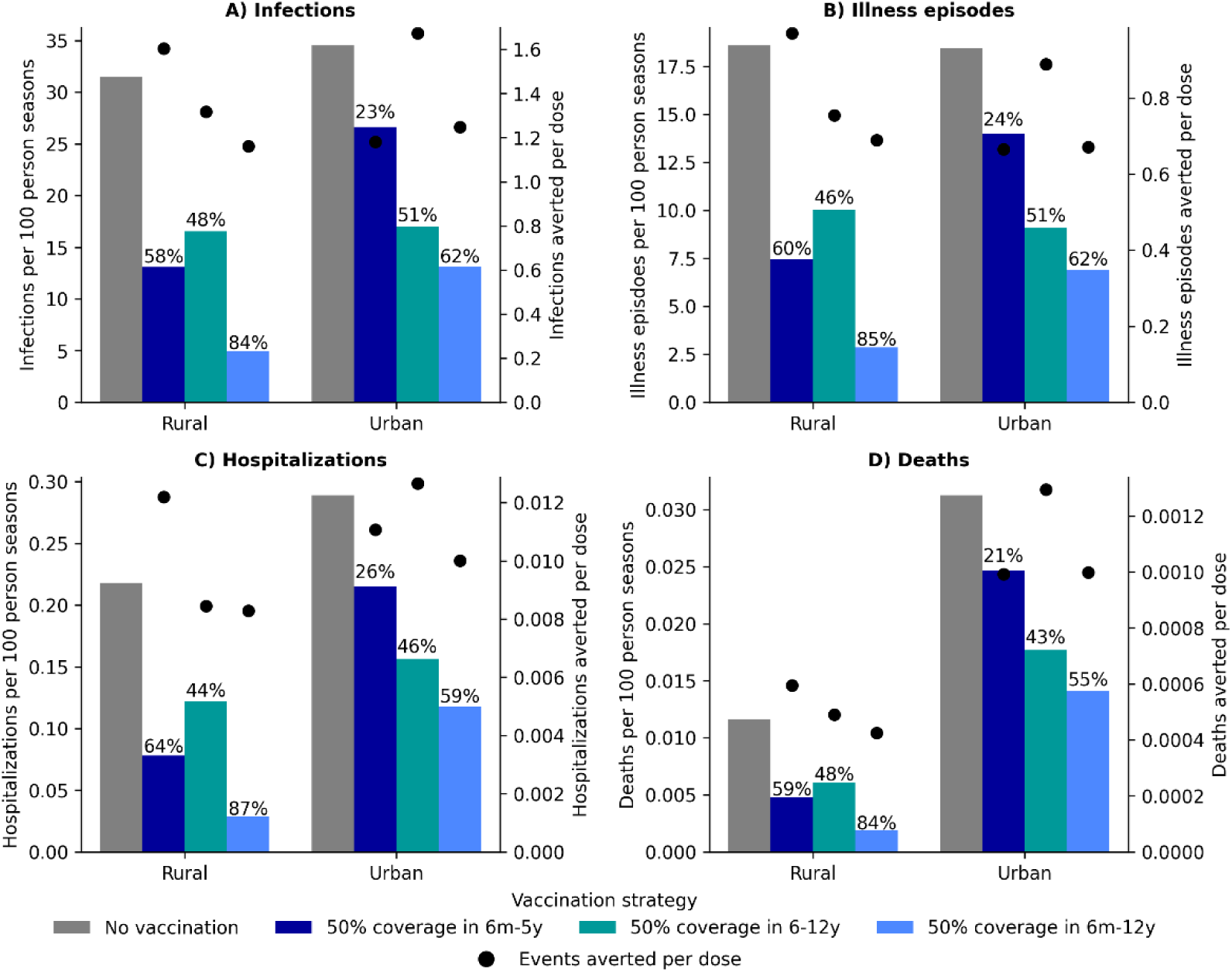
Annual average projected burden of influenza and reductions due to vaccinations. A) infections, B) illness episodes (symptomatic illness), C) hospitalizations and D) deaths per 100 person seasons (bars, primary axis). Results are shown for the base model (no vaccination) and three scenarios of vaccinating 50% of the target age group (6 months to 5 years, 6-12 years and 6 months – 12 years) with a vaccine with 50% effectiveness against infection and duration of shedding. The graph displays annual event rates (left y-axis, infections, illness episodes, hospitalizations or deaths), and the number of averted events per vaccine dose (dots, secondary axis). Percentage reduction is also shown (text at edge of bar). Data are based on model projections for urban and rural sites, averaged over the 2016-2018 seasons, South Africa

## Discussion

By modeling high-resolution data on infection, serology, and contacts from a South African household cohort study, we evaluated the potential benefits of pediatric influenza vaccination programs in mitigating community-wide transmission and disease burden. Our modeling analysis indicates that even with moderate vaccination coverage (50%) and modest vaccine efficacy against both infection and transmission (50% each, respectively), a transmission-reducing immunization campaign targeting children could yield substantial benefits. Specifically, such a strategy may reduce infections, illness episodes, hospitalizations, and deaths by 21–87% in an average season, depending on circulating strain, geographical settings, and targeted age groups (Figure 4).

Even without accounting for the effects of vaccine strain match and the timing of influenza vaccination relative to the influenza season, we observed considerable differences in the overall reduction of influenza incidence across seasons and sites. These variations were likely driven by a combination of factors, including the circulating subtype/lineage and age-based attack rates for the specific season, as well as the population age structure and contact patterns. Here we only considered how variability in influenza attack rates may affect the population-level benefits of vaccination, but additional variation in VE have been reported between seasons and populations due to vaccine mismatch and immunologic factors [27]. Our data highlights how the optimal pediatric influenza vaccine program may vary across seasons and sites, driven by different mean ages at infection by season and geography.

Our cohort study in South Africa revealed very high attack rates of influenza infection in the population (44 infections per 100 person seasons), with rates as high as 67 infections per 100 person seasons in children younger than 5 years [4]. Children also shed virus for longer and at higher viral loads [4, 28–30], which translates to higher transmission potential [4, 21, 30–32]. Taken together, these factors make a pediatric influenza vaccine program particularly attractive for South Africa, as demonstrated by our model projections. Previous modeling studies have similarly demonstrated promising direct and indirect benefits of pediatric vaccination. For example, modeling of next-generation influenza vaccines in Kenya demonstrated both direct and indirect effects of pediatric programs, with a 29% to 66% reduction in infections in all ages depending on the product used, achieving cost-effectiveness [12]. These results align well with estimates from our models. A model from the Netherlands also showed pediatric influenza vaccination to have large indirect effects, with almost half of the symptomatic cases and almost all deaths averted arising from the non-vaccinated age groups [10]. A United Kingdom (UK) model showed lower reductions on infections but higher reductions in deaths when vaccinating 50% of children aged 6 months to 4 years compared to our model (0.5 infections averted per dose and 1.8 deaths averted per 1,000 doses in UK model vs 1.2 infections averted per dose and 0.4 deaths averted per dose in our model) [11]. Differences between the two studies are likely due to the difference in age structure between the two settings, the UK having less children (hence less infections and presumably less indirect effects) and a greater population aged ≥65 years (hence more deaths per capita) [33].

Further evidence for the impact of pediatric influenza vaccine programs also comes from observational studies of large-scale vaccine rollouts. The UK extended their influenza vaccination program from the elderly to children aged 6 months to 17 years starting in 2013. Results from comparing primary school pilot to non-pilot areas showed a 59% reduction in general practitioner influenza-like illness consultations in the 2014/15 season [19], although no significant reductions were observed in the 2015/16 season [20]. Estimation of vaccine effects from observational ecological studies of time trends is notoriously difficult, and impact estimates may differ considerably between observational studies and transmission models. A US study of school-based vaccination programs revealed notable differences between the two approaches, with observational approaches estimating much higher indirect effects (33%) than mathematical models (2%). These discrepancies between empirical and modeling studies highlight the need to integrate real-world data with mathematical approaches to improve benefits estimates and address potential biases [13].

The assumption of a vaccine that can reduce 50% of onward transmission by shortening the duration of shedding is not based on a currently available product. For mRNA COVID-19 vaccines, it has been demonstrated that vaccination provides indirect protection to close contacts [34–36], as well as reduces onward transmission from breakthrough infections [35]. Data are scarce on the indirect effects of vaccinating children for influenza. A community-based randomized control trial investigating the direct and indirect effects of inactivated influenza vaccine (IIV) showed only a 20% reduction in infection probability for household members if a single child aged 5-17 years in the household is vaccinated [37]. On the other hand, vaccinating children in daycare with IIV resulted in 42% fewer respiratory illnesses in household contacts [15]. A cluster randomized control trial in which children aged 3-15 years were vaccinated IIV showed a 61% protection against influenza infection in non-vaccinated community members [14]. Specifically, due to the induction of mucosal immunity, live attenuated influenza vaccine (LAIV) has greater potential to reduce onward transmission. A cluster randomized trial of elementary schools where schools were allocated to receive either IIV or LAIV showed a 64% greater protection in household members against influenza infection when a child was vaccinated with LAIV compared to IIV [16].

With new influenza vaccine products on the horizon, including >200 universal vaccine candidates on different virus, nucleic acid, virus-like particles (VLP), non-VLP nanoparticles, recombinant protein, and virus vector platforms [38], it is important to understand the key targets and endpoints that future vaccines should aim for to achieve the greatest impact on transmission and overall disease burden. Our model framework is adaptable to various VE estimates, including reductions in susceptibility, duration and peak of viral shedding, making it a versatile tool to assess and optimize new influenza vaccine products as they become available.

A key strength of our model stems from robust parameterization using comprehensive data from multi-year cohorts, providing detailed characterization of influenza transmission dynamics, including pre-season antibody titers, symptomatic fraction, duration of shedding, risk of infection and transmission [4, 24], as well as contact rates within these communities [25]. These data enabled us to significantly reduce the number of parameters that need to be estimated, while reproducing the overall temporal and age dynamics of influenza. In addition, use of a detailed model allowed us to simulate different mechanisms of vaccine protection.

A few limitations are worth noting. Firstly, our model was focused on the dynamics of influenza within a single season, and we did not consider waning immunity and viral evolution that may affect multi-year influenza dynamics. We implicitly accounted for inter-annual fluctuations in population immunity by initializing each season based on pre-existing titers to circulating strains in the cohort. We also estimated subtype/lineage-specific transmission rates each season, allowing for differences in force-of-infection between seasons and subtype/lineage. Our model was already highly complex and it would have been difficult to explicitly account for additional mechanisms such as influenza inter-annual variability, especially given the unknown duration of protection following infection.

Secondly, while our model was calibrated to data from two South African communities, and may not be generalizable to other countries, we believe our results provide reasonable estimates for communities with similar population structures and contact patterns. Thirdly, recognizing that HAI titers are only one aspect of the immune response to influenza, we did not incorporate the boosting of HAI titers following vaccination or its potential impact on vaccine efficacy. Instead, we applied a pre-specified vaccine efficacy, defined as a percentage reduction in both shedding duration and susceptibility. Lastly, our analysis primarily focused on the evaluation of a vaccine campaign’s indirect effects. We thus ignored the direct effects of vaccination in further reducing mild to severe disease among vaccine breakthrough infections, particularly because the campaign was targeted at children, who generally have a lower risk of severe illness. As a result, our projections of vaccine benefits are likely conservative.

In conclusion, our study provides compelling evidence that pediatric influenza immunization programs employing transmission-reducing vaccines can substantially lower the community burden of influenza, particularly in settings with high attack rates among children. Our model offers a valuable tool for determining the minimum efficacy requirement for targeted direct and indirect impact on influenza burden, as well as for evaluating vaccine impact with established efficacy profiles. From a translational perspective, it can inform clinical trial design by assisting in defining target product profiles to achieve a predetermined population impact, and by guiding the development of post-licensure immunization programs to maximize benefits during large-scale rollouts.

## Data Availability

All data produced in the present work are contained in the manuscript. Model code will be made available after publication in peer-review journal.

## Acknowledgement

This work was supported by the National Institute for Communicable Diseases of the National Health Laboratory Service and the US Centers for Disease Control and Prevention (cooperative agreement number 5U51IP000155), and the in-house research program of the Division of International Epidemiology and Population Studies, Fogarty International Center, National Institutes of Health.

The findings and conclusions in this paper are those of the authors and do not necessarily represent the official position of the NICD, NIH, US CDC. CC has received grant support from Sanofi Pasteur, US CDC, Welcome Trust, Taskforce for Global Health and Bill & Melinda Gates Foundation. AvG has received grant support from US CDC, ASLM/Africa CDC and WHO Afro. NW reports receiving grants from Sanofi Pasteur, US CDC and the Gates Foundation. JM has received grant support from Sanofi Pasteur and PATH. Other authors declare no competing interests.

## Author contributions

### Conception and design of the study

JK, CV, ST, CC, KS

### Data collection and laboratory processing

JK, ST, NW, JM, AvG, LM, AM, CC

### Analysis and interpretation

JK, CV, MS, ST, NW, JM, AvG, LM, AM, CC, KS

### Drafting or critical review of the article

JK, CV, MS, ST, NW, JM, AvG, LM, AM, CC, KS

### Data reporting

Model code is available at https://github.com/crdm-nicd/phirst_fluvax_transm_model. [*Will be made available upon publication*]

## Supplementary files

Supplementary Excel file 1: Model parameter values. Supplementary tables and figures.

---

**Fig. S1:**
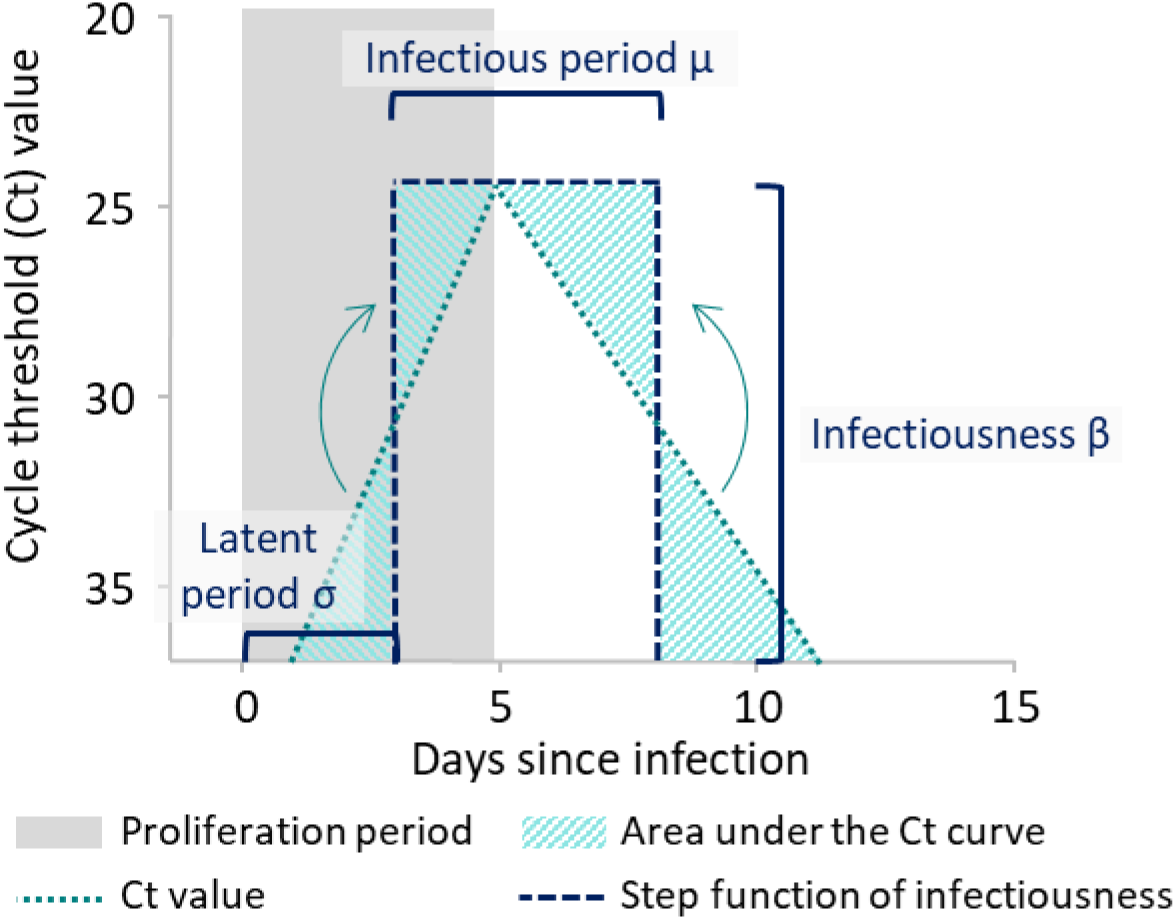
Definition of latent period, infectious period and infectiousness. Estimated within age and pre-season HAI titer.

**Fig. S2:**
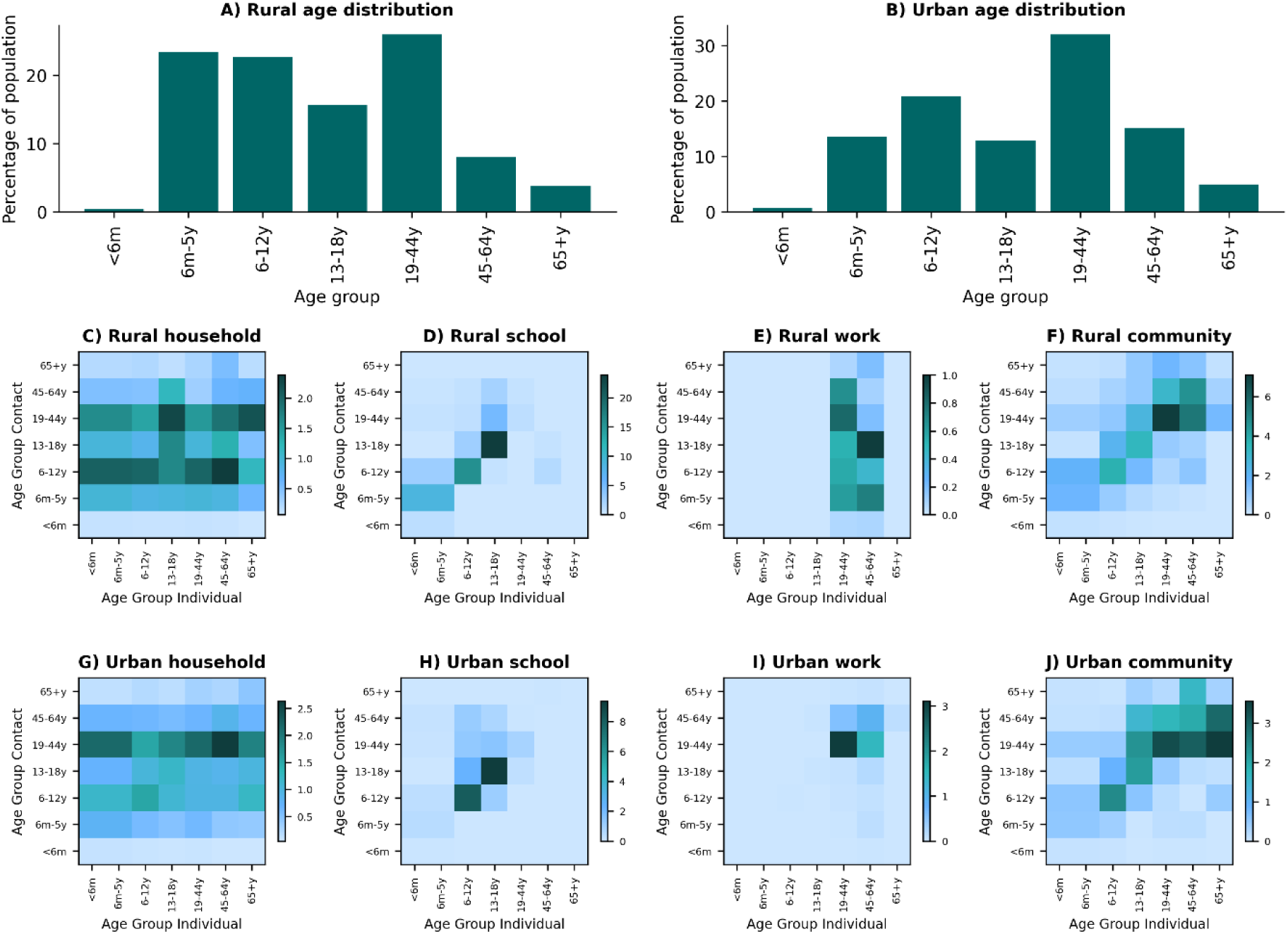
Age distribution at the A) rural and B) urban sites. Average number of contacts in the household (C, G), school (D, H), work (E, I) and community (F, J) at the rural (C-F) and urban (G-J) sites, 2018, South Africa

**Fig. S3:**
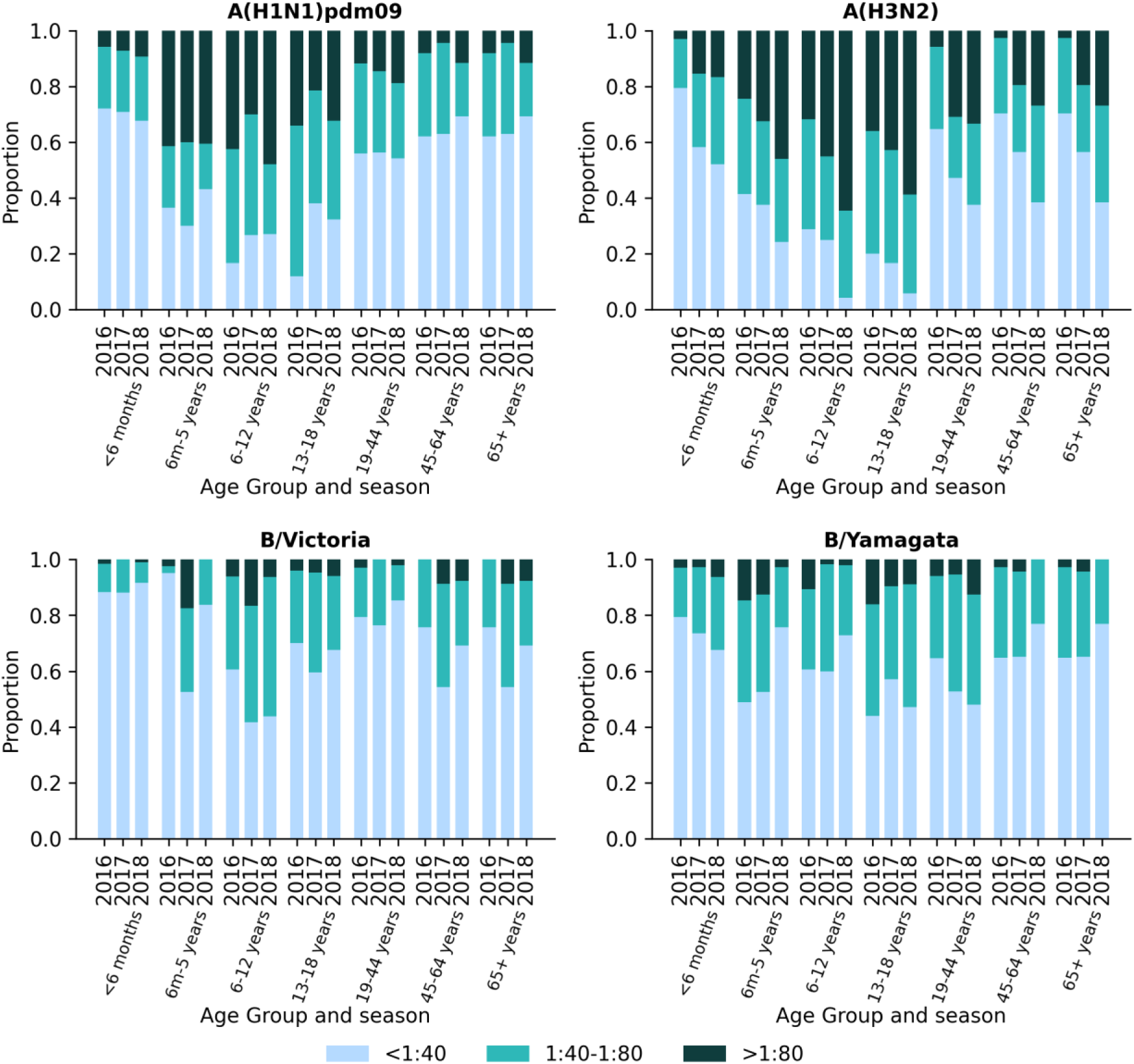
Proportion of population with pre-season hemagglutination inhibition assay (HAI) titers for A(H1N1)pdm09, A(H3N2), B/Victoria and B/Yamagata within each age group and season, rural site, 2016-2018, South Africa

**Fig. S4:**
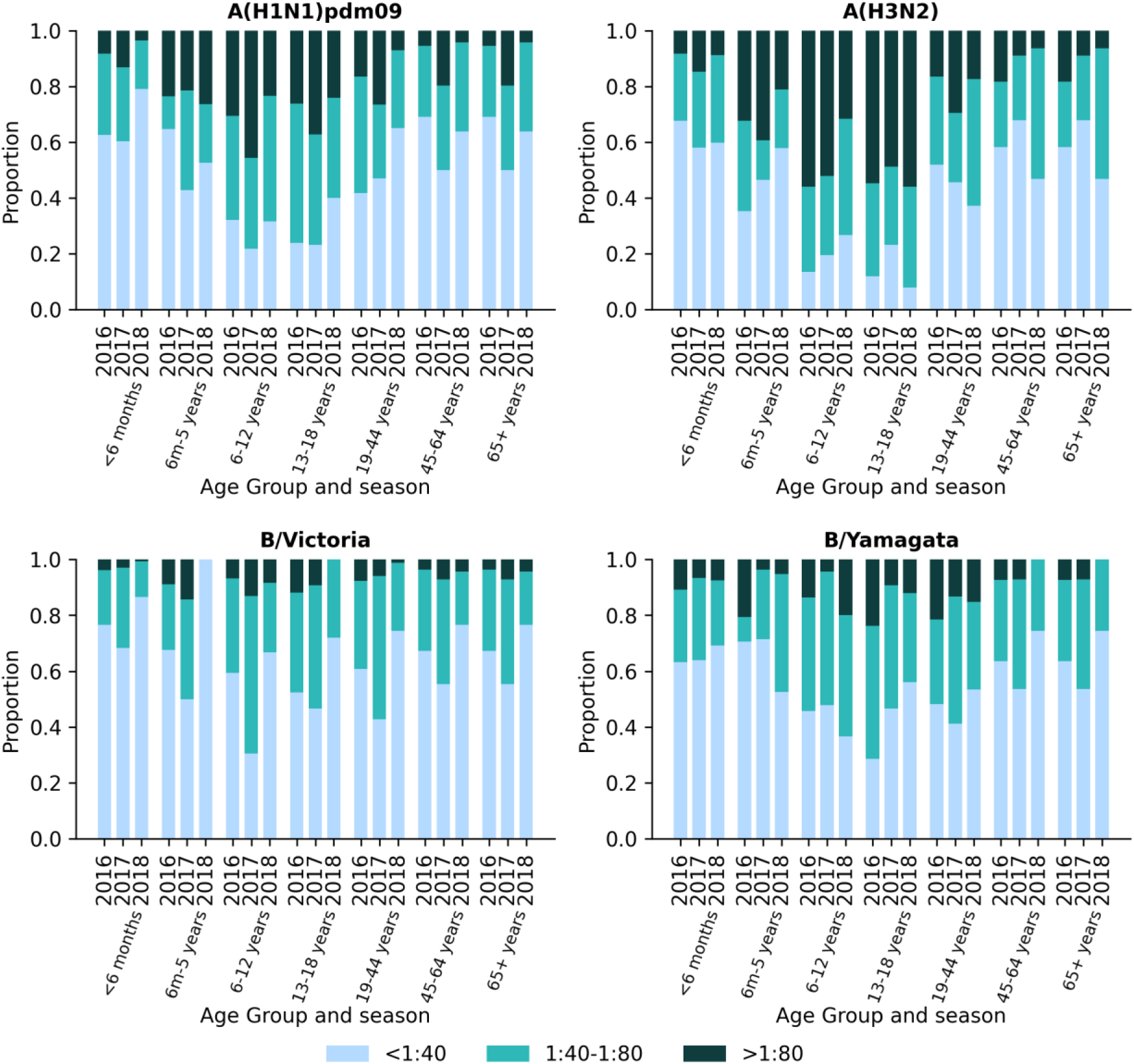
Proportion of population with pre-season hemagglutination inhibition assay (HAI) titers for A(H1N1)pdm09, A(H3N2), B/Victoria and B/Yamagata within each age group and season, urban site, 2016-2018, South Africa

**Fig. S5:**
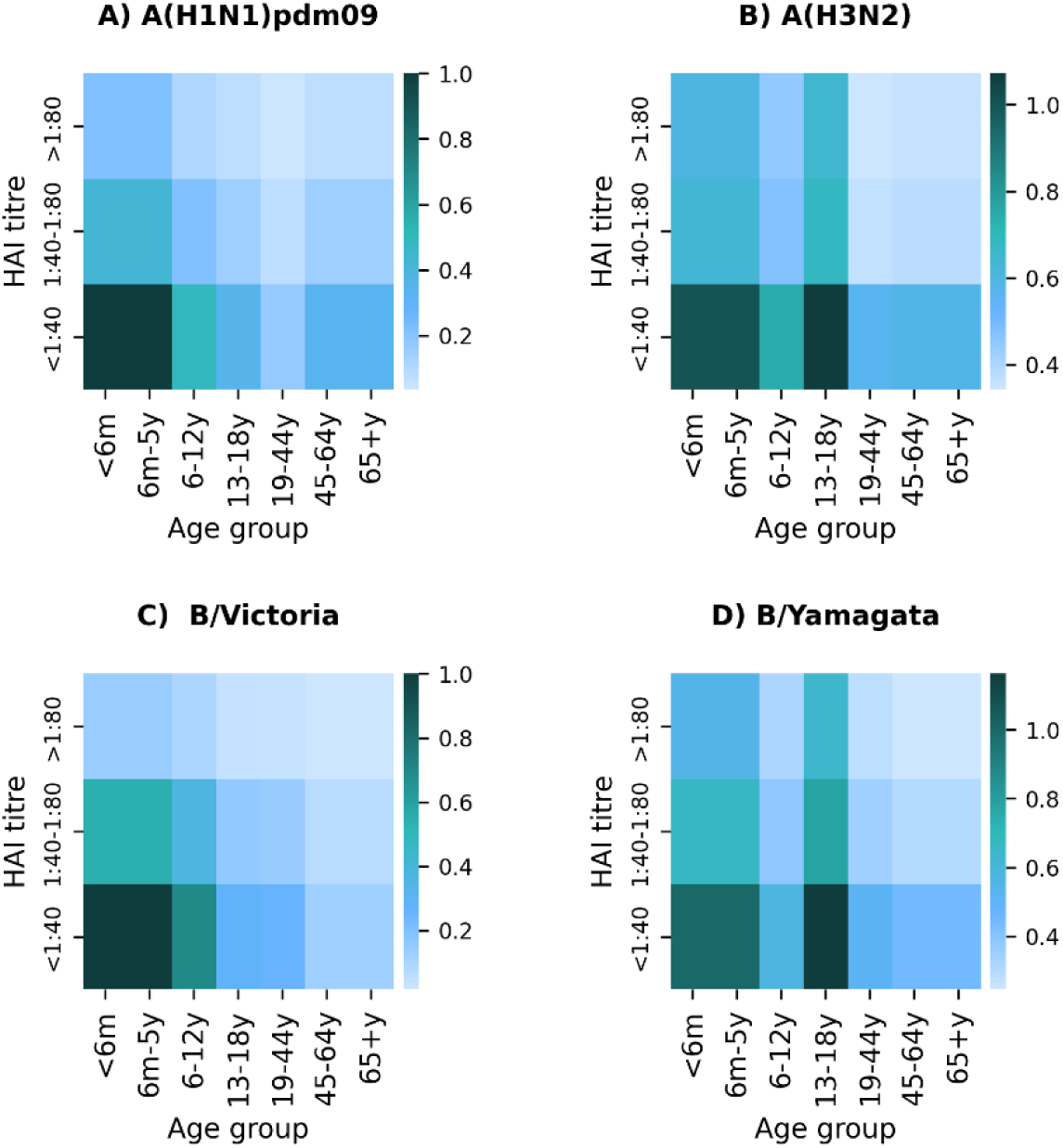
Susceptibility (α, risk of infection) by age and pre-season hemagglutination inhibition assay (HAI) titers for A(H1N1)pdm09, A(H3N2), B/Victoria and B/Yamagata, both sites, 2016-2018, South Africa

**Fig. S6:**
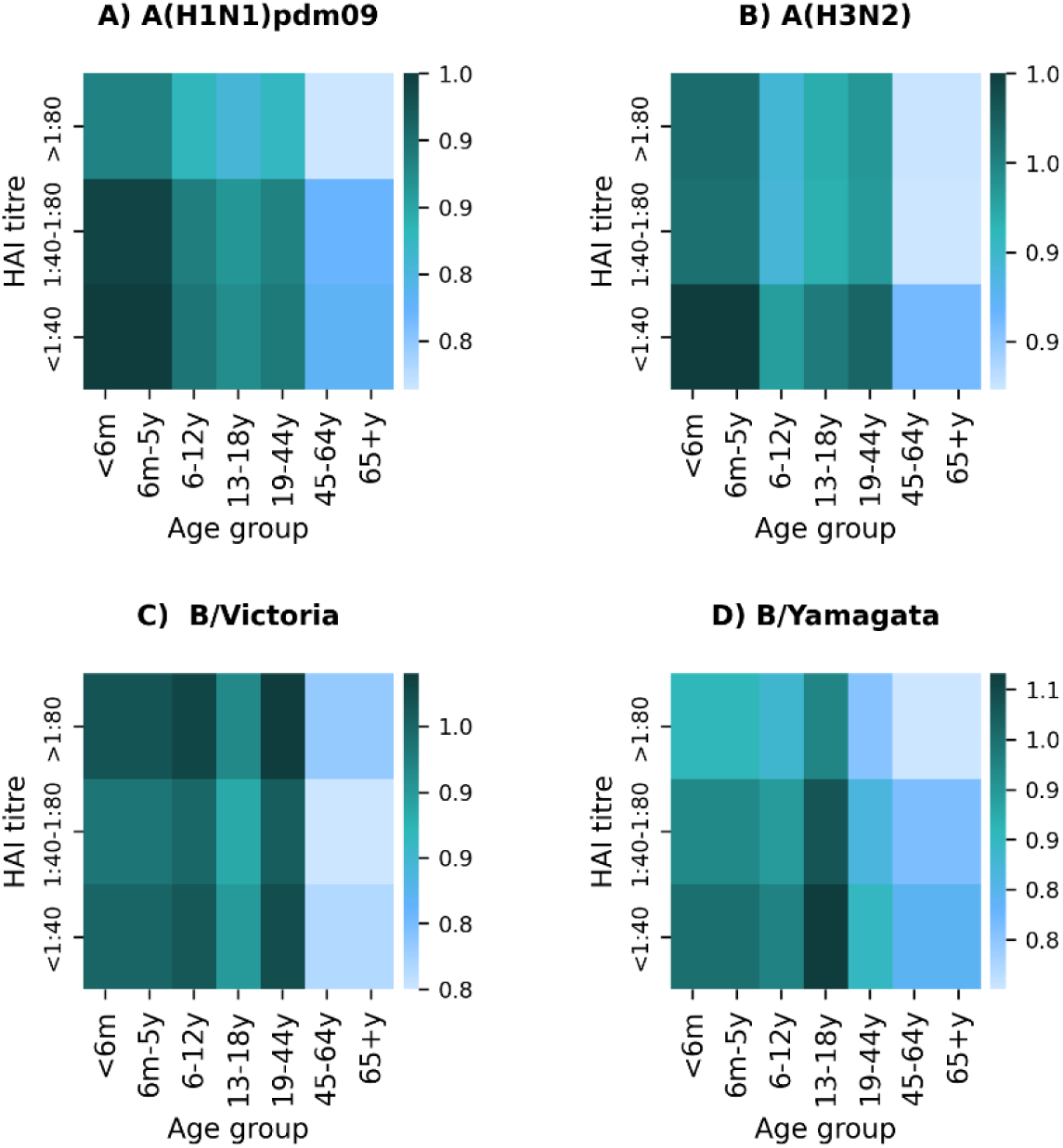
Infectiousness (β, minimum Ct value as proxy for viral load) by age and pre-season hemagglutination inhibition assay (HAI) titers for A(H1N1)pdm09, A(H3N2), B/Victoria and B/Yamagata, both sites, 2016-2018, South Africa

**Fig. S7:**
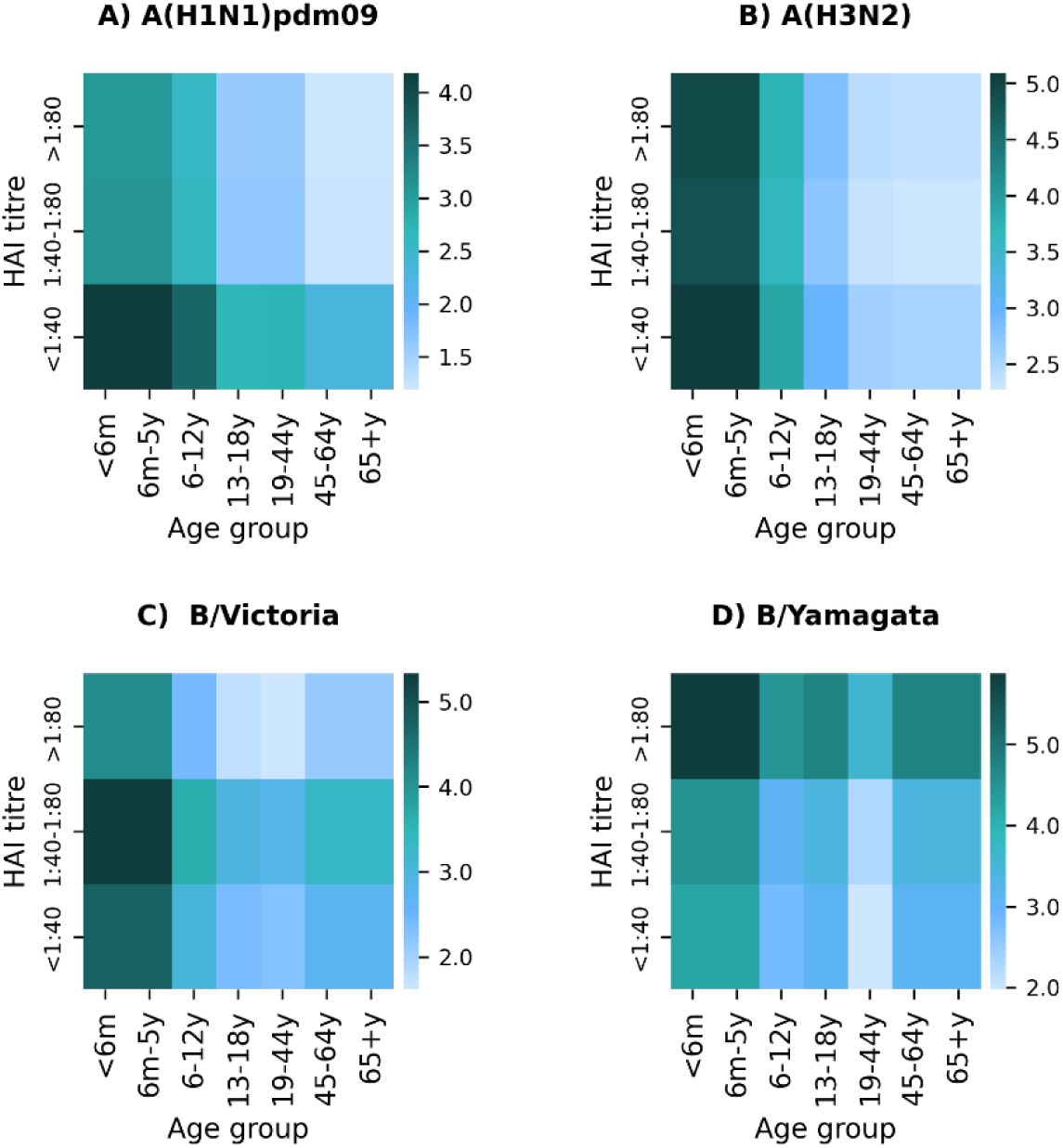
Infectious period (µ, ½ duration of shedding) by age and pre-season hemagglutination inhibition assay (HAI) titers for A(H1N1)pdm09, A(H3N2), B/Victoria and B/Yamagata, both sites, 2016-2018, South Africa

**Fig. S8:**
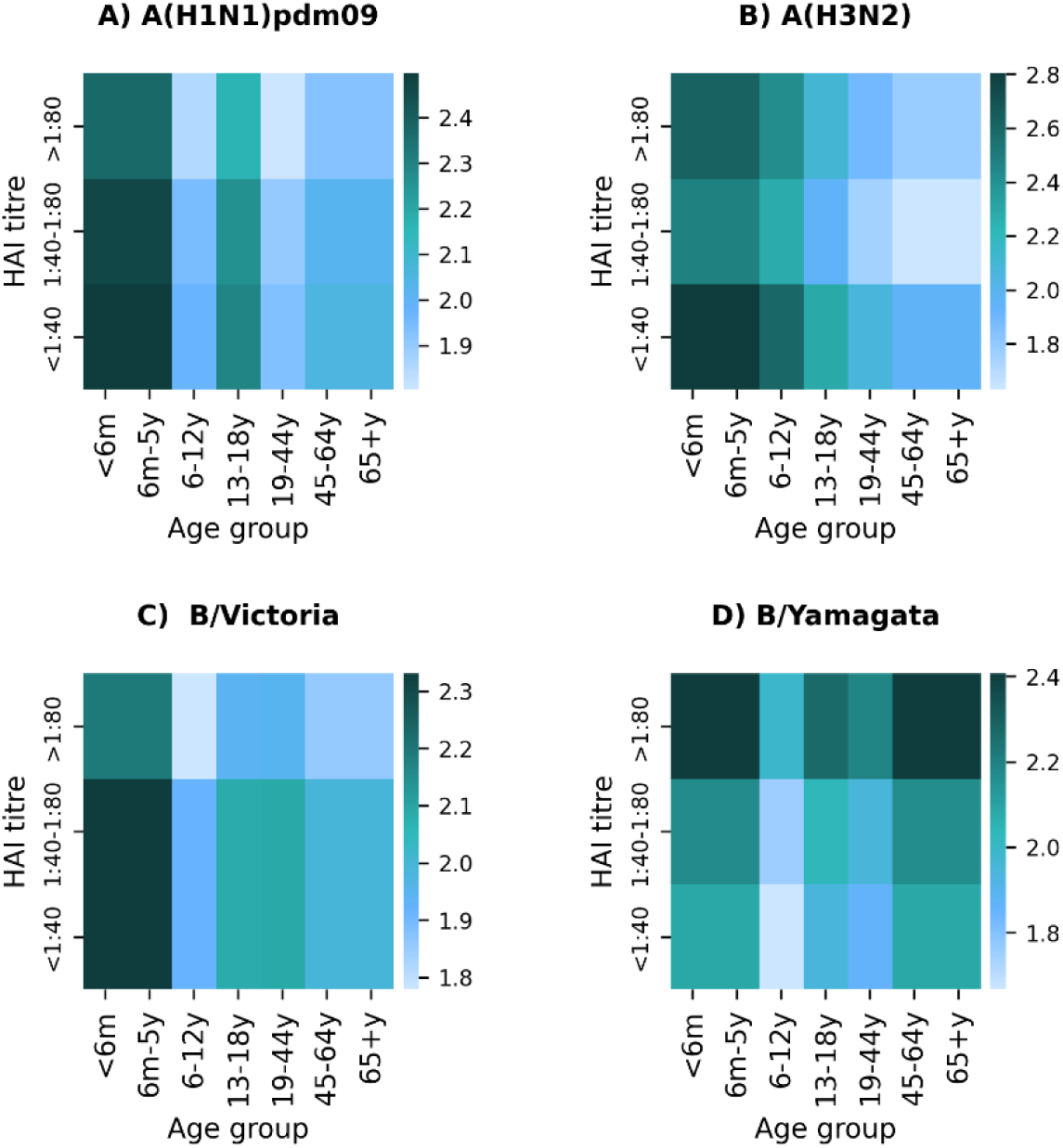
Latent period (σ, infection to infectiousness) by age and pre-season hemagglutination inhibition assay (HAI) titers for A(H1N1)pdm09, A(H3N2), B/Victoria and B/Yamagata, both sites, 2016-2018, South Africa

**Fig. S9:**
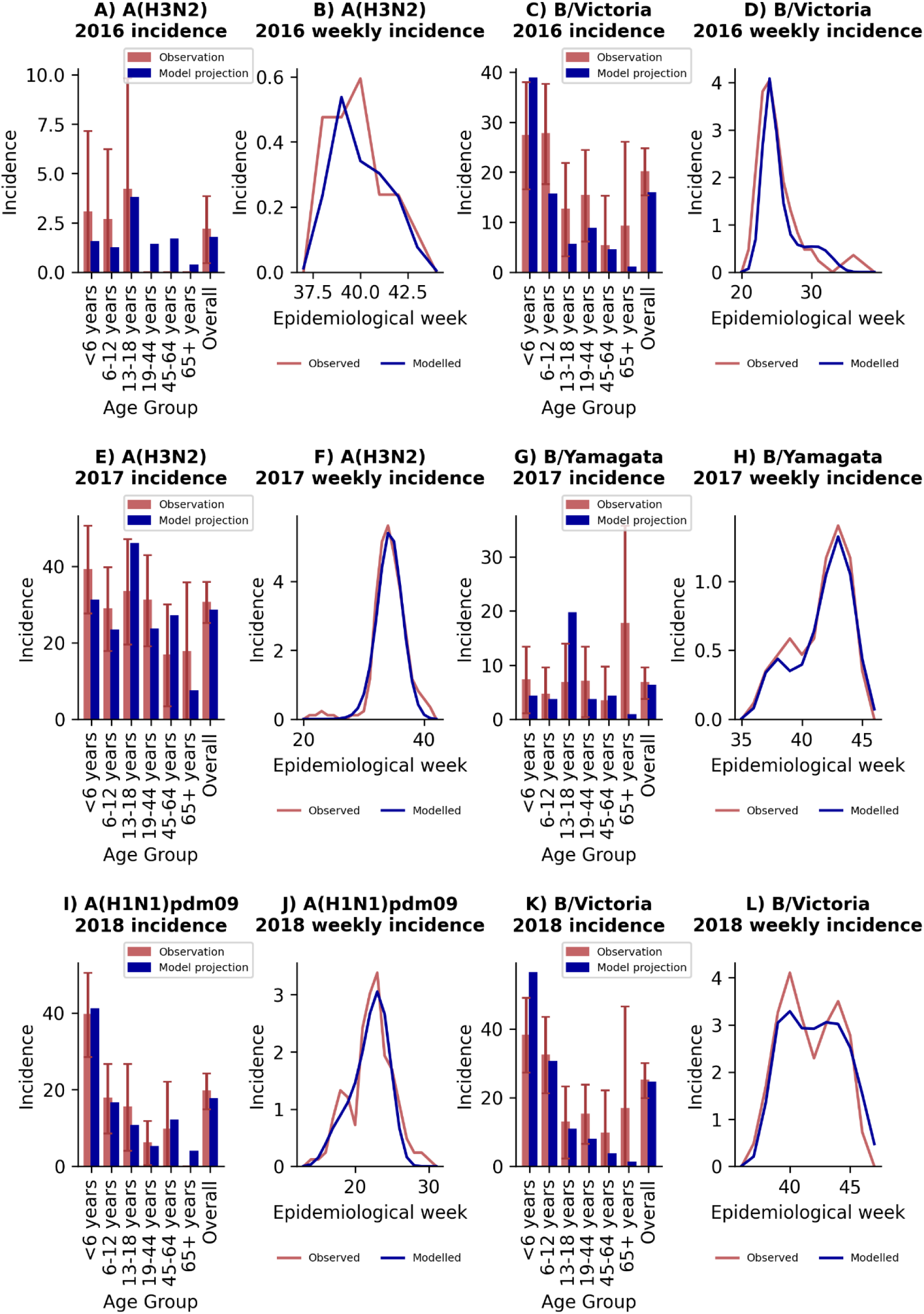
Observed and modelled incidence per 100 population (with 95% confidence intervals) by age group and weekly observed and modelled incidence used for model calibration for each season (2016 (A-D), 2017 (E-H), 2018 (I-L) and subtype/lineage, rural site, South Africa.

**Fig. S10:**
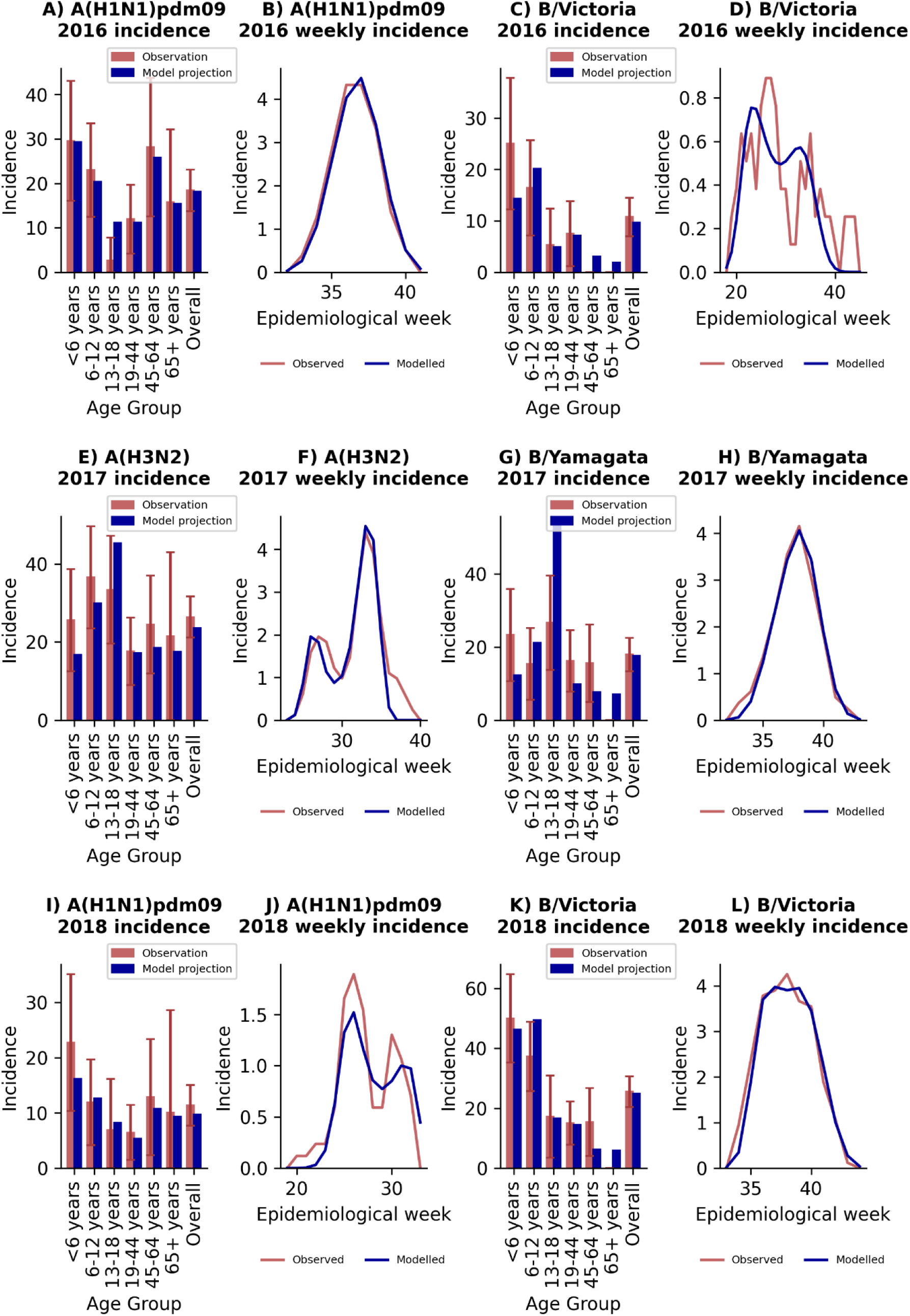
Observed and modelled incidence per 100 population (with 95% confidence intervals) by age group and weekly observed and modelled incidence per 100 population used for model calibration for each season (2016 (A-D), 2017 (E-H), 2018 (I-L) and subtype/lineage, urban site, South Africa.

**Fig. S11:**
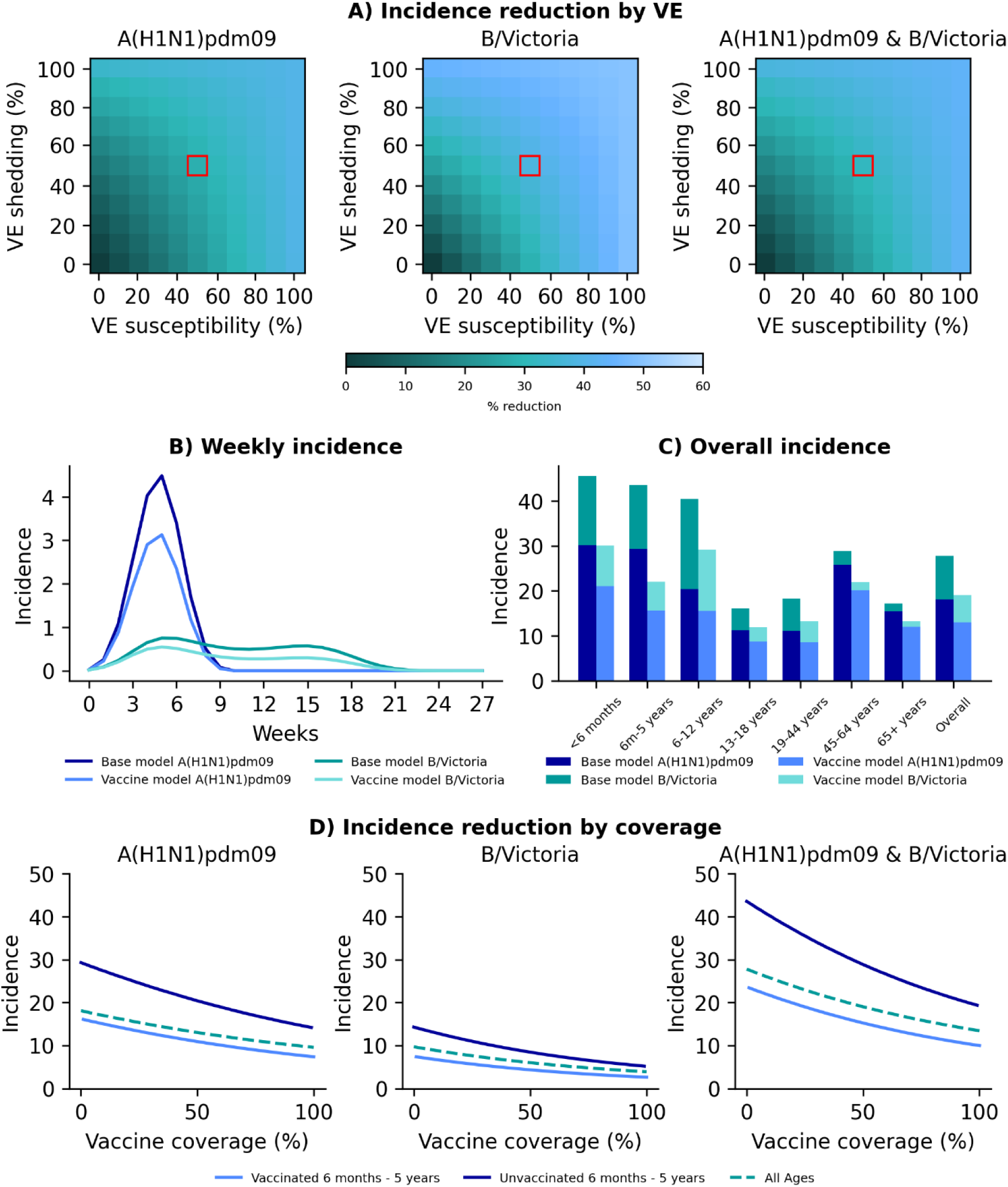
Vaccine model results for vaccinating children aged 6 months - 5 years, urban site, 2016 season. A) Percentage reduction in incidence at differing levels of vaccine effectiveness (VE) against susceptibility (x axis) and duration of shedding (y axis) when vaccinating 50% of the target age group. Red block denotes VE chosen (50%) for subsequent analysis. B) Modelled weekly incidence of individual influenza subtype/lineage in base model (no vaccination) and vaccine model. C) Overall incidence of influenza subtype/lineage by age in base and vaccine model. Both B and C based on 50% coverage in target age group with vaccine with VE of 50% against susceptibility and duration of shedding. D) Incidence of influenza subtype/lineage (left and middle) and combined season (right) at varying vaccine coverage levels in the target age group with vaccine with VE of 50% against susceptibility and duration of shedding.

**Fig. S12:**
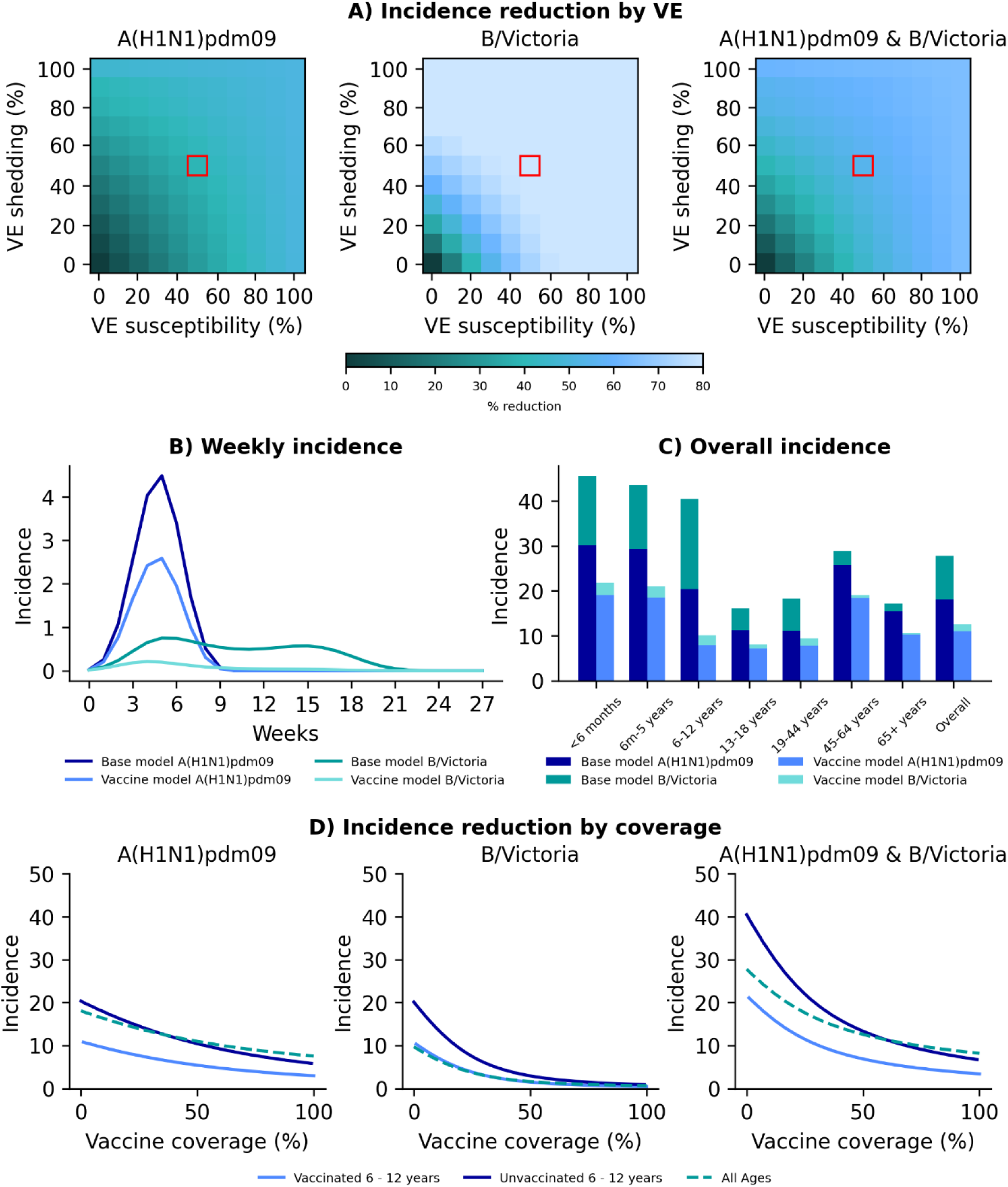
Vaccine model results for vaccinating children aged 6 - 12 years, urban site, 2016 season. A) Percentage reduction in incidence at differing levels of vaccine effectiveness (VE) against susceptibility (x axis) and duration of shedding (y axis) when vaccinating 50% of the target age group. Red block denotes VE chosen (50%) for subsequent analysis. B) Modelled weekly incidence per 100 population of individual influenza subtype/lineage in base model (no vaccination) and vaccine model. C) Overall incidence of influenza per 100 population subtype/lineage by age in base and vaccine model. Both B and C based on 50% coverage in target age group with vaccine with VE of 50% against susceptibility and duration of shedding. D) Incidence of influenza per 100 population subtype/lineage (left and middle) and combined season (right) at varying vaccine coverage levels in the target age group with vaccine with VE of 50% against susceptibility and duration of shedding.

**Fig. S13:**
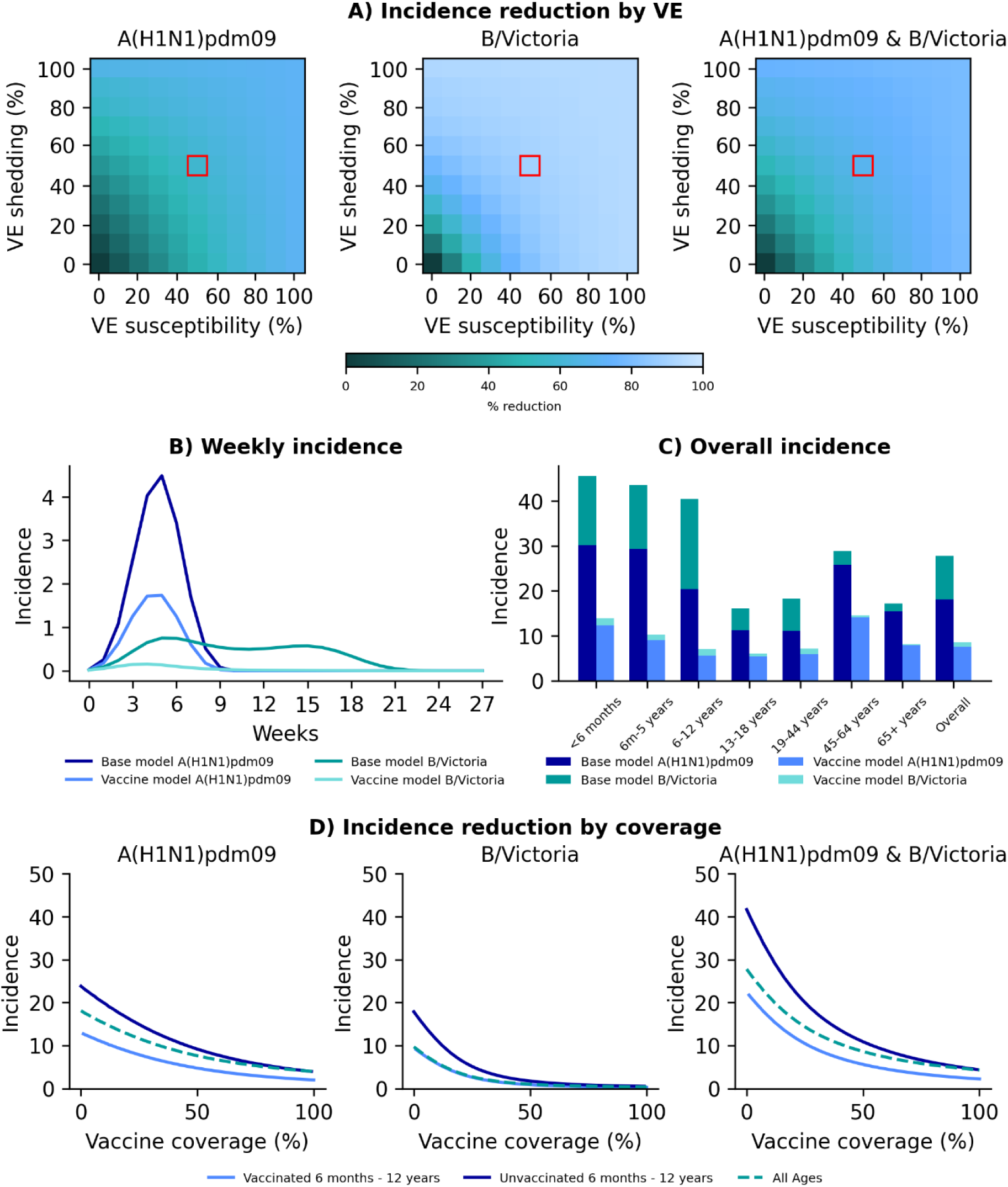
Vaccine model results for vaccinating children aged 6 months - 12 years, urban site, 2016 season. A) Percentage reduction in incidence at differing levels of vaccine effectiveness (VE) against susceptibility (x axis) and duration of shedding (y axis) when vaccinating 50% of the target age group. Red block denotes VE chosen (50%) for subsequent analysis. B) Modelled weekly incidence per 100 population of individual influenza subtype/lineage in base model (no vaccination) and vaccine model. C) Overall incidence of influenza per 100 population subtype/lineage by age in base and vaccine model. Both B and C based on 50% coverage in target age group with vaccine with VE of 50% against susceptibility and duration of shedding. D) Incidence of influenza per 100 population subtype/lineage (left and middle) and combined season (right) at varying vaccine coverage levels in the target age group with vaccine with VE of 50% against susceptibility and duration of shedding.

**Fig. S14:**
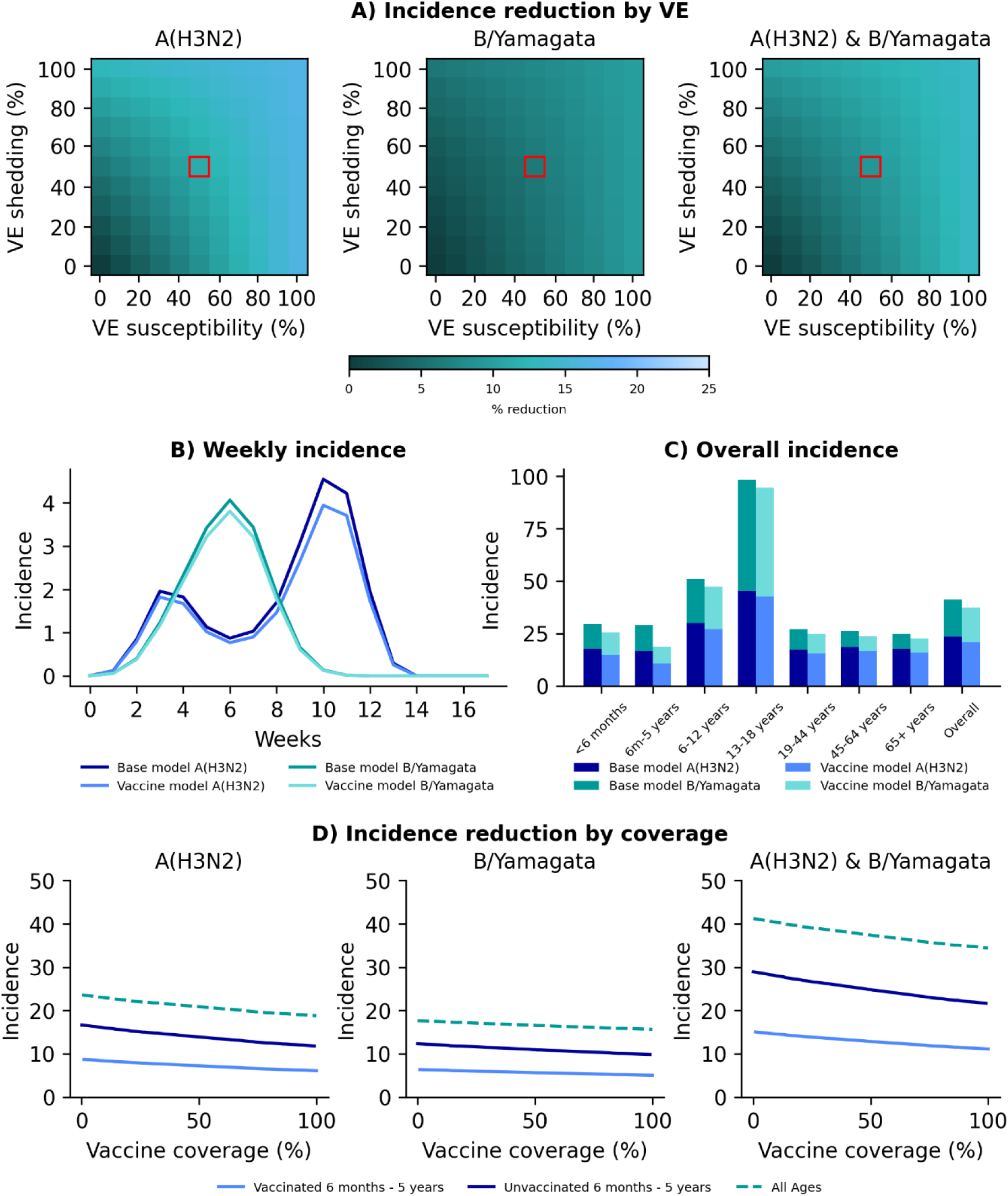
Vaccine model results for vaccinating children aged 6 months - 5 years, urban site, 2017 season. A) Percentage reduction in incidence at differing levels of vaccine effectiveness (VE) against susceptibility (x axis) and duration of shedding (y axis) when vaccinating 50% of the target age group. Red block denotes VE chosen (50%) for subsequent analysis. B) Modelled weekly incidence per 100 population of individual influenza subtype/lineage in base model (no vaccination) and vaccine model. C) Overall incidence of influenza per 100 population subtype/lineage by age in base and vaccine model. Both B and C based on 50% coverage in target age group with vaccine with VE of 50% against susceptibility and duration of shedding. D) Incidence of influenza per 100 population subtype/lineage (left and middle) and combined season (right) at varying vaccine coverage levels in the target age group with vaccine with VE of 50% against susceptibility and duration of shedding.

**Fig. S15:**
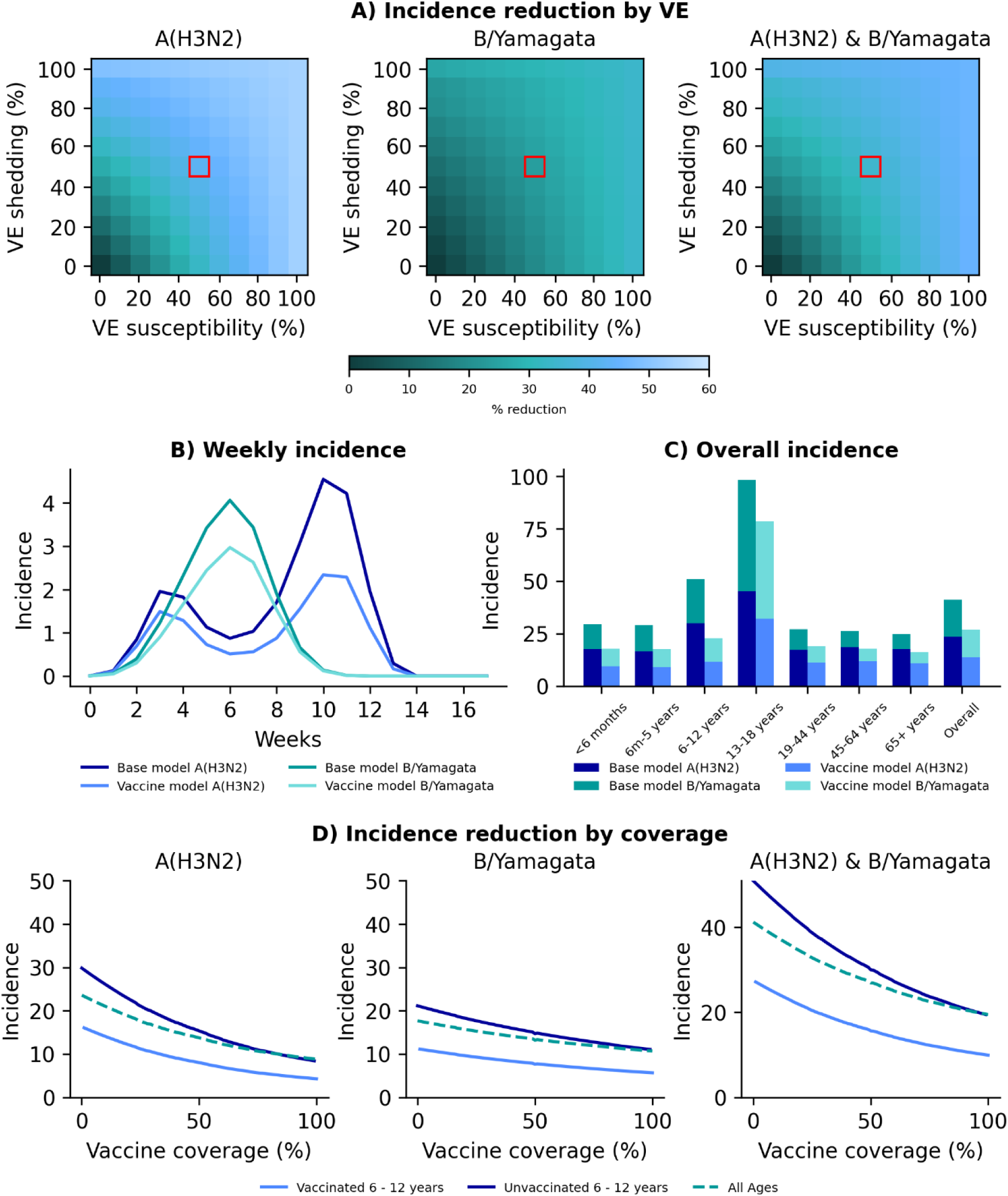
Vaccine model results for vaccinating children aged 6 - 12 years, urban site, 2017 season. A) Percentage reduction in incidence at differing levels of vaccine effectiveness (VE) against susceptibility (x axis) and duration of shedding (y axis) when vaccinating 50% of the target age group. Red block denotes VE chosen (50%) for subsequent analysis. B) Modelled weekly incidence per 100 population of individual influenza subtype/lineage in base model (no vaccination) and vaccine model. C) Overall incidence of influenza per 100 population subtype/lineage by age in base and vaccine model. Both B and C based on 50% coverage in target age group with vaccine with VE of 50% against susceptibility and duration of shedding. D) Incidence of influenza per 100 population subtype/lineage (left and middle) and combined season (right) at varying vaccine coverage levels in the target age group with vaccine with VE of 50% against susceptibility and duration of shedding.

**Fig. S16:**
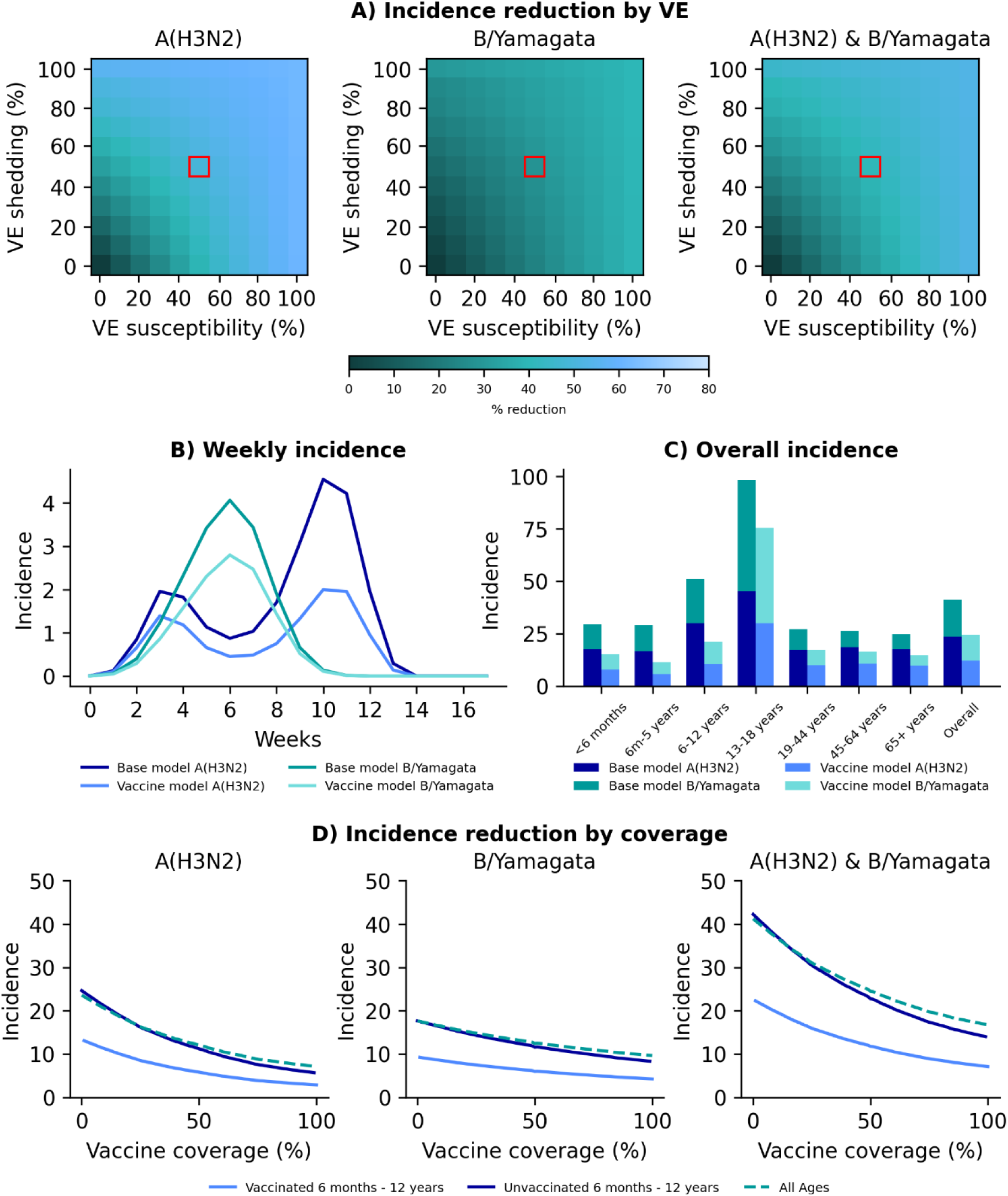
Vaccine model results for vaccinating children aged 6 months - 12 years, urban site, 2017 season. A) Percentage reduction in incidence at differing levels of vaccine effectiveness (VE) against susceptibility (x axis) and duration of shedding (y axis) when vaccinating 50% of the target age group. Red block denotes VE chosen (50%) for subsequent analysis. B) Modelled weekly incidence per 100 population of individual influenza subtype/lineage in base model (no vaccination) and vaccine model. C) Overall incidence of influenza per 100 population subtype/lineage by age in base and vaccine model. Both B and C based on 50% coverage in target age group with vaccine with VE of 50% against susceptibility and duration of shedding. D) Incidence of influenza per 100 population subtype/lineage (left and middle) and combined season (right) at varying vaccine coverage levels in the target age group with vaccine with VE of 50% against susceptibility and duration of shedding.

**Fig. S17:**
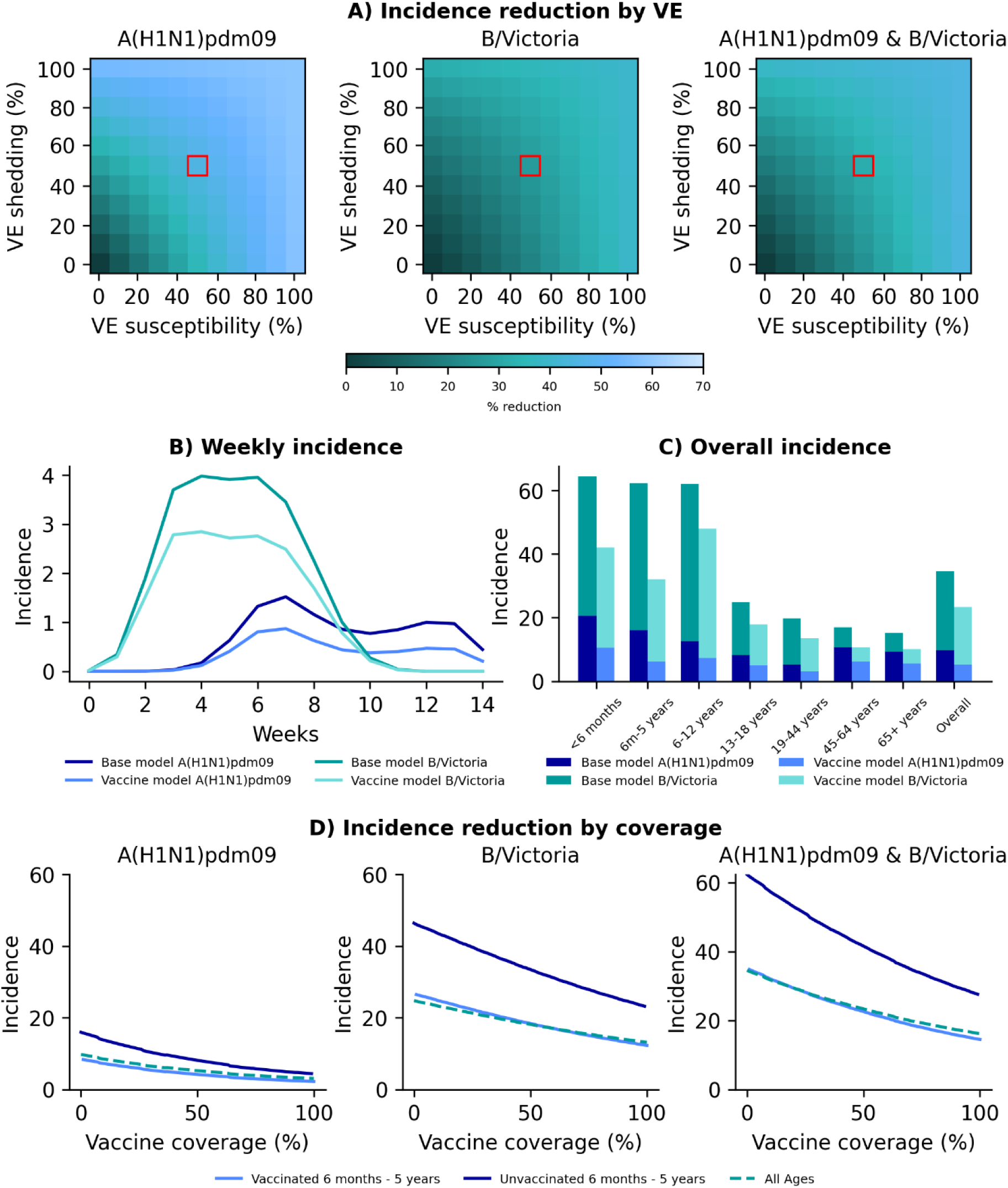
Vaccine model results for vaccinating children aged 6 months - 5 years, urban site, 2018 season. A) Percentage reduction in incidence at differing levels of vaccine effectiveness (VE) against susceptibility (x axis) and duration of shedding (y axis) when vaccinating 50% of the target age group. Red block denotes VE chosen (50%) for subsequent analysis. B) Modelled weekly incidence per 100 population of individual influenza subtype/lineage in base model (no vaccination) and vaccine model. C) Overall incidence of influenza per 100 population subtype/lineage by age in base and vaccine model. Both B and C based on 50% coverage in target age group with vaccine with VE of 50% against susceptibility and duration of shedding. D) Incidence of influenza per 100 population subtype/lineage (left and middle) and combined season (right) at varying vaccine coverage levels in the target age group with vaccine with VE of 50% against susceptibility and duration of shedding.

**Fig. S18:**
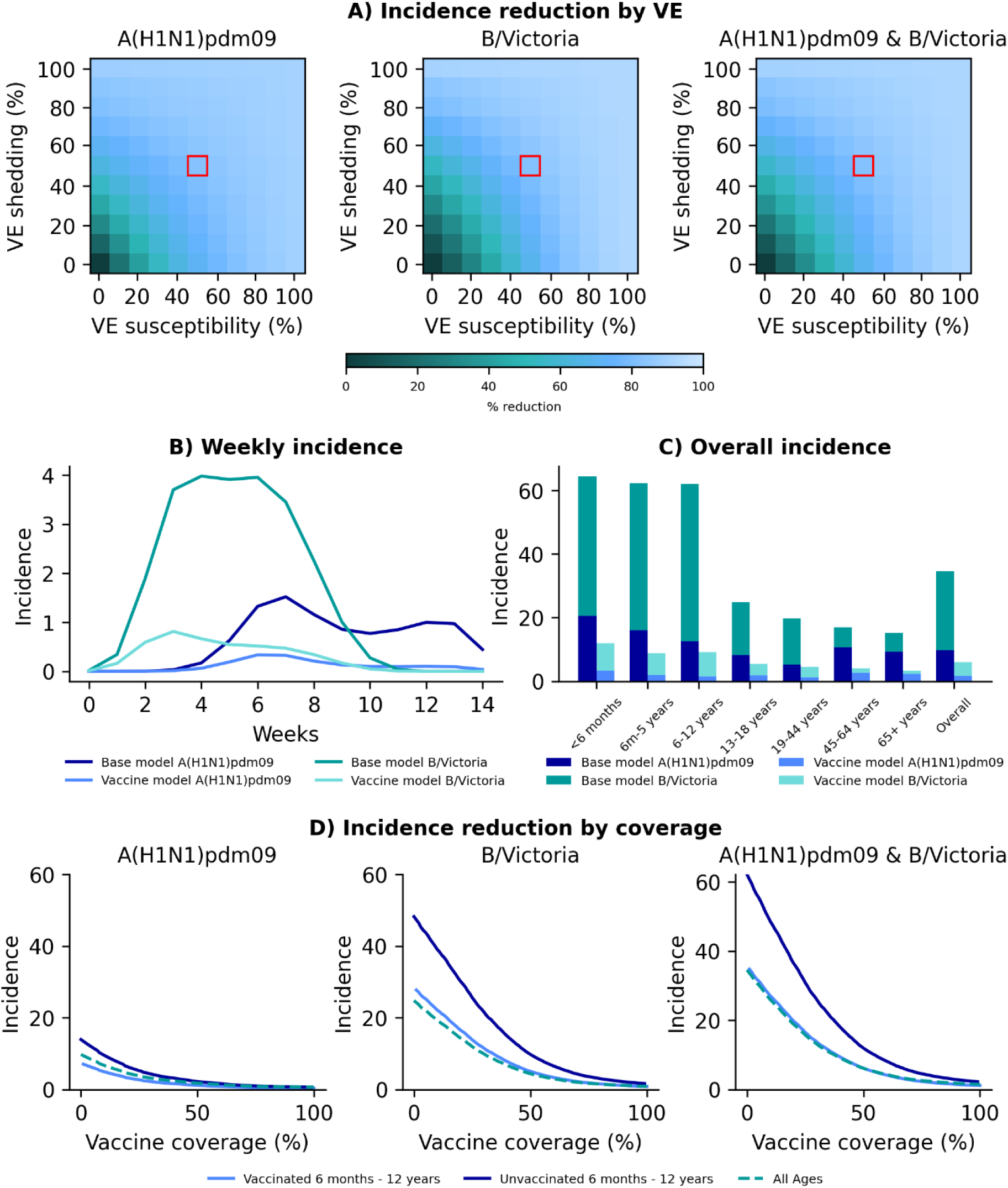
Vaccine model results for vaccinating children aged 6 - 12 years, urban site, 2018 season. A) Percentage reduction in incidence at differing levels of vaccine effectiveness (VE) against susceptibility (x axis) and duration of shedding (y axis) when vaccinating 50% of the target age group. Red block denotes VE chosen (50%) for subsequent analysis. B) Modelled weekly incidence per 100 population of individual influenza subtype/lineage in base model (no vaccination) and vaccine model. C) Overall incidence of influenza per 100 population subtype/lineage by age in base and vaccine model. Both B and C based on 50% coverage in target age group with vaccine with VE of 50% against susceptibility and duration of shedding. D) Incidence of influenza per 100 population subtype/lineage (left and middle) and combined season (right) at varying vaccine coverage levels in the target age group with vaccine with VE of 50% against susceptibility and duration of shedding.

**Fig. S19:**
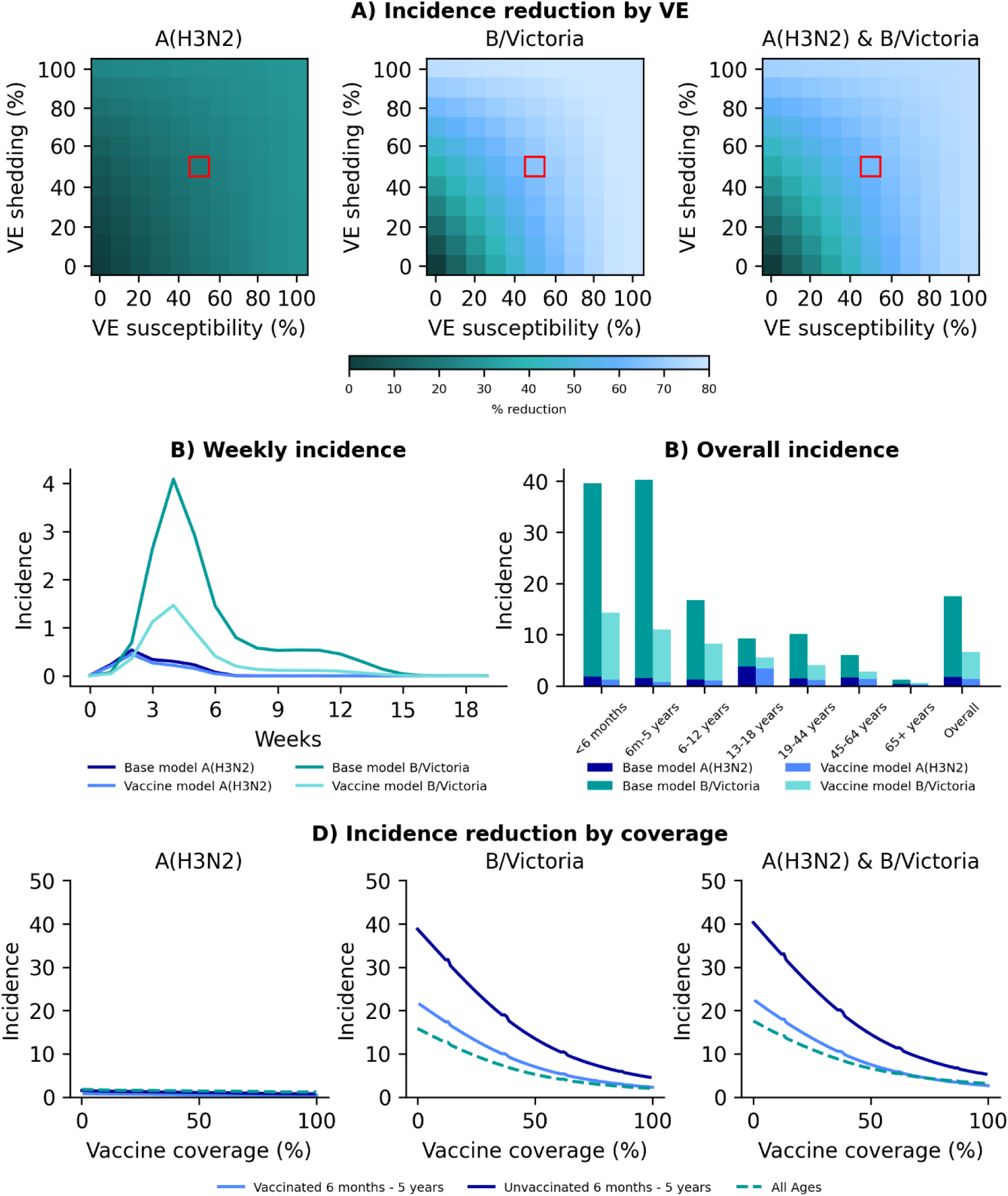
Vaccine model results for vaccinating children aged 6 months - 5 years, rural site, 2016 season. A) Percentage reduction in incidence at differing levels of vaccine effectiveness (VE) against susceptibility (x axis) and duration of shedding (y axis) when vaccinating 50% of the target age group. Red block denotes VE chosen (50%) for subsequent analysis. B) Modelled weekly incidence per 100 population of individual influenza subtype/lineage in base model (no vaccination) and vaccine model. C) Overall incidence of influenza per 100 population subtype/lineage by age in base and vaccine model. Both B and C based on 50% coverage in target age group with vaccine with VE of 50% against susceptibility and duration of shedding. D) Incidence of influenza per 100 population subtype/lineage (left and middle) and combined season (right) at varying vaccine coverage levels in the target age group with vaccine with VE of 50% against susceptibility and duration of shedding.

**Fig. S20:**
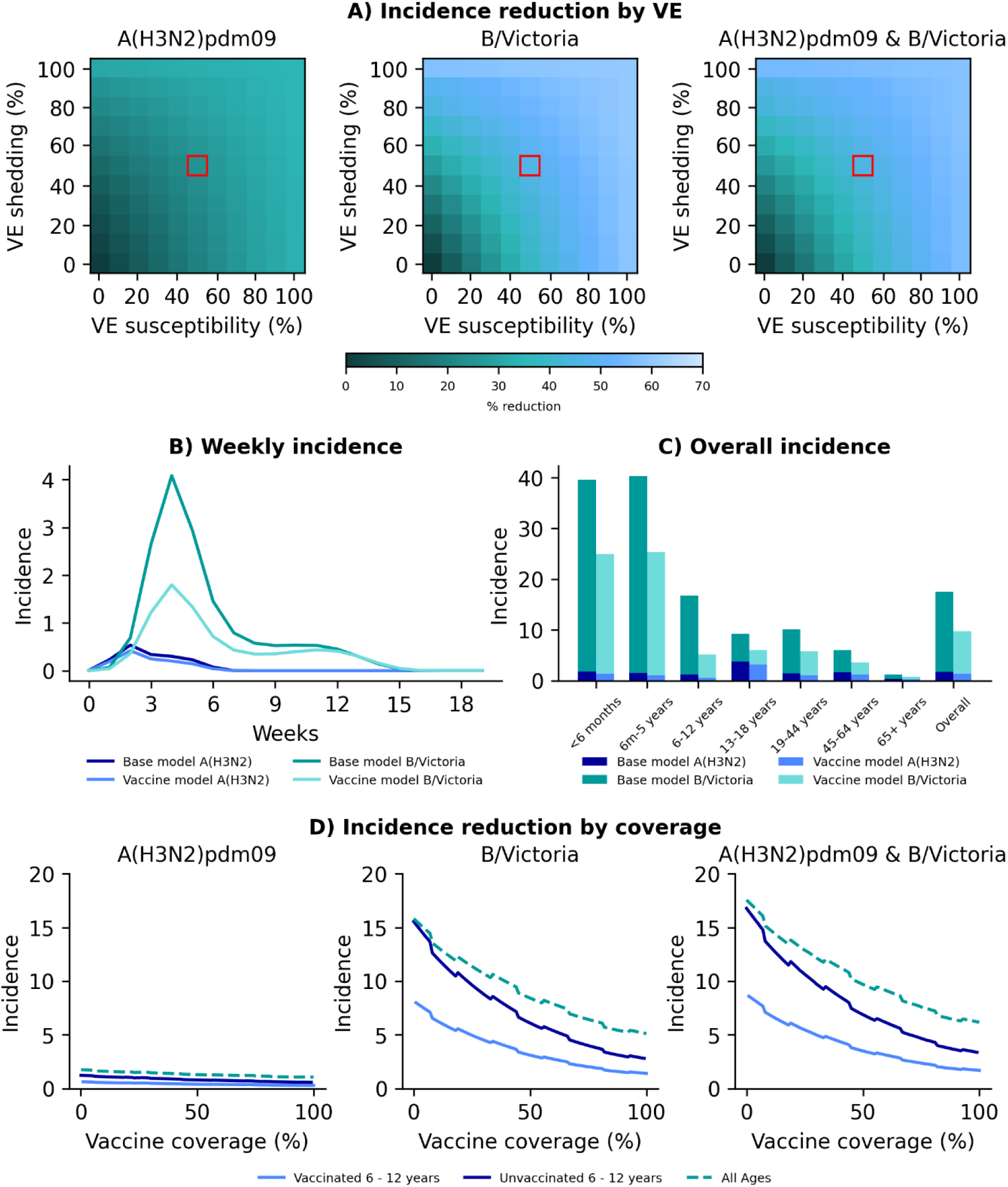
Vaccine model results for vaccinating children aged 6 - 12 years, rural site, 2016 season. A) Percentage reduction in incidence at differing levels of vaccine effectiveness (VE) against susceptibility (x axis) and duration of shedding (y axis) when vaccinating 50% of the target age group. Red block denotes VE chosen (50%) for subsequent analysis. B) Modelled weekly incidence per 100 population of individual influenza subtype/lineage in base model (no vaccination) and vaccine model. C) Overall incidence of influenza per 100 population subtype/lineage by age in base and vaccine model. Both B and C based on 50% coverage in target age group with vaccine with VE of 50% against susceptibility and duration of shedding. D) Incidence of influenza per 100 population subtype/lineage (left and middle) and combined season (right) at varying vaccine coverage levels in the target age group with vaccine with VE of 50% against susceptibility and duration of shedding.

**Fig. S21:**
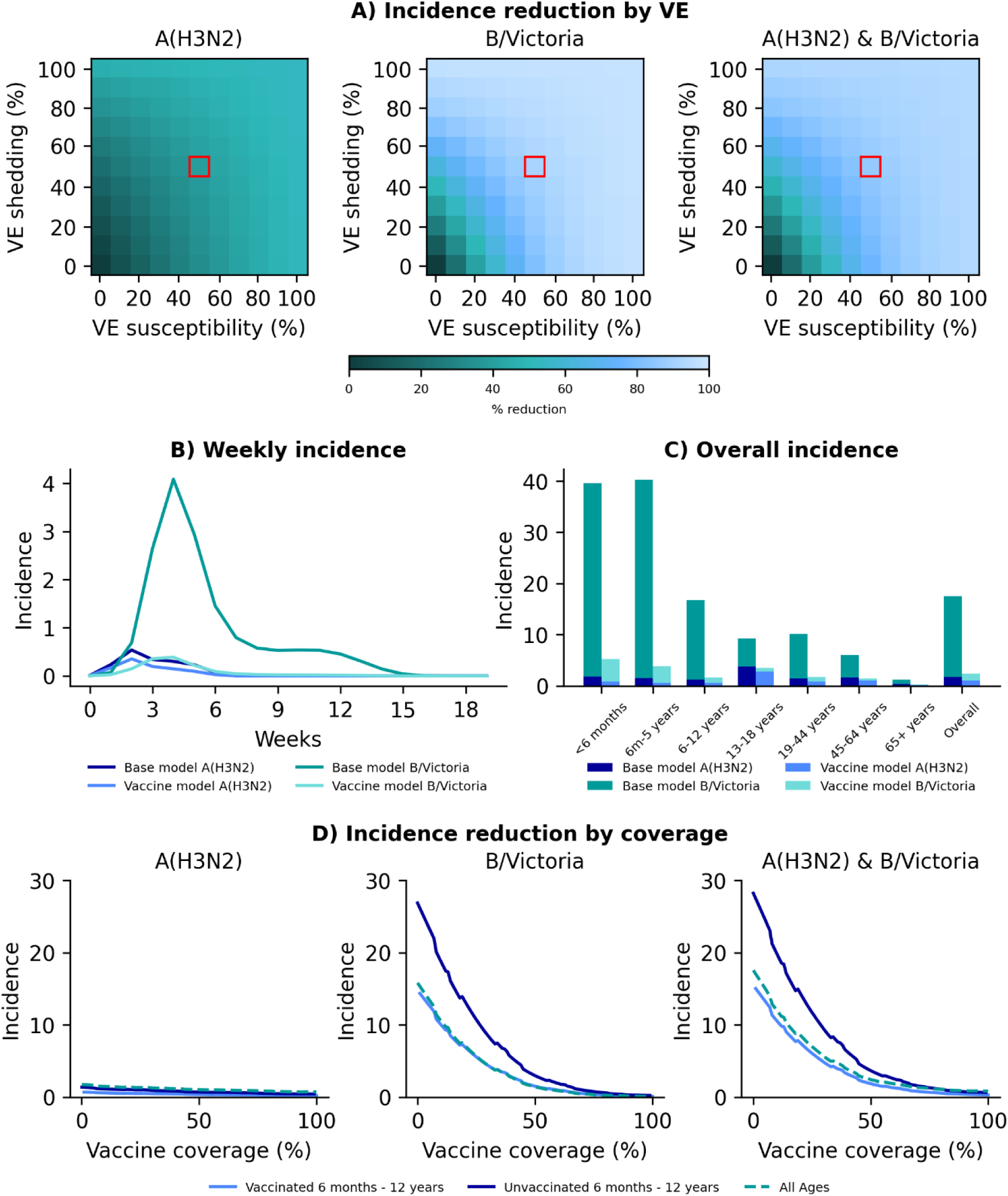
Vaccine model results for vaccinating children aged 6 months - 12 years, rural site, 2016 season. A) Percentage reduction in incidence at differing levels of vaccine effectiveness (VE) against susceptibility (x axis) and duration of shedding (y axis) when vaccinating 50% of the target age group. Red block denotes VE chosen (50%) for subsequent analysis. B) Modelled weekly incidence per 100 population of individual influenza subtype/lineage in base model (no vaccination) and vaccine model. C) Overall incidence of influenza per 100 population subtype/lineage by age in base and vaccine model. Both B and C based on 50% coverage in target age group with vaccine with VE of 50% against susceptibility and duration of shedding. D) Incidence of influenza per 100 population subtype/lineage (left and middle) and combined season (right) at varying vaccine coverage levels in the target age group with vaccine with VE of 50% against susceptibility and duration of shedding.

**Fig. S22:**
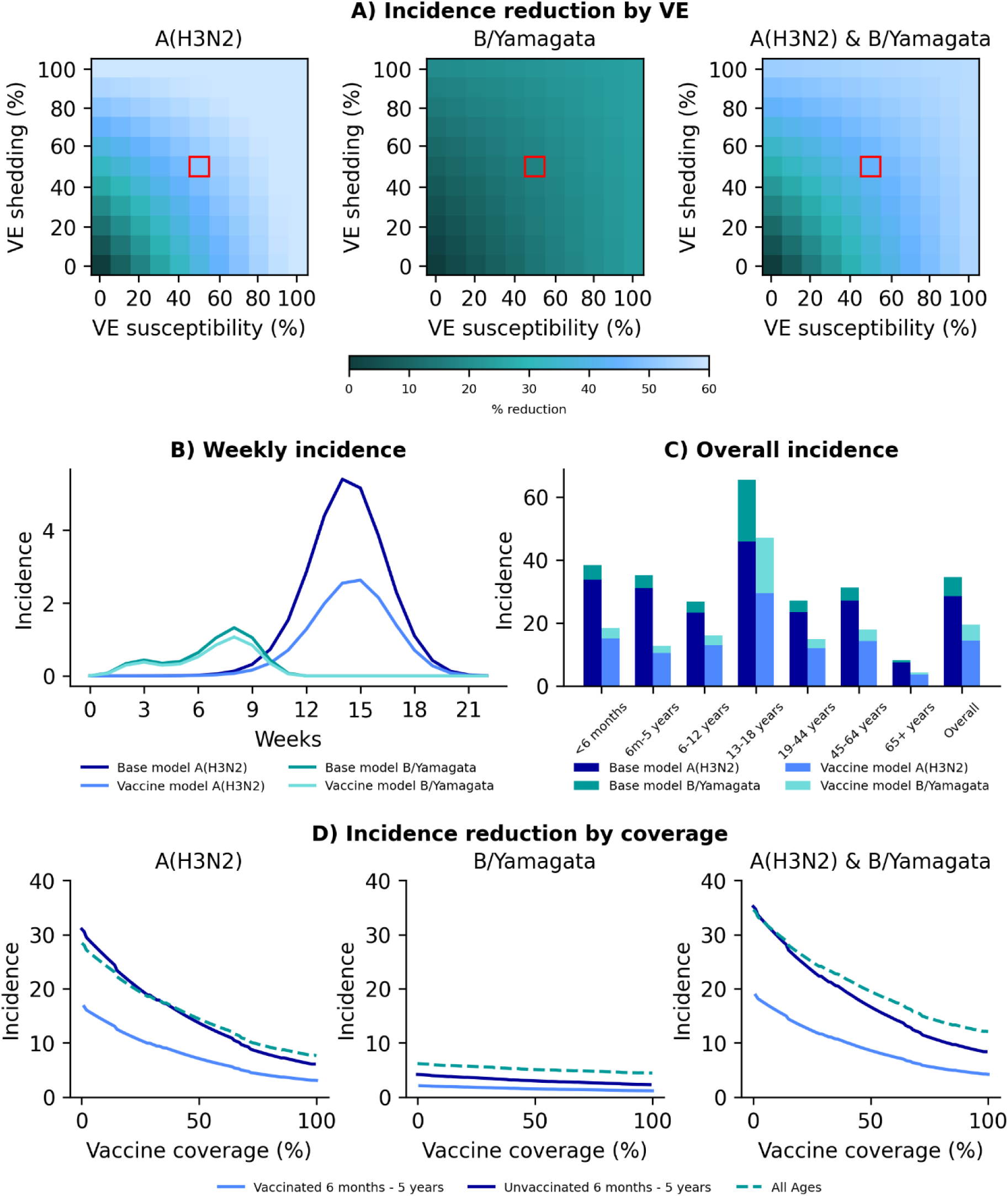
Vaccine model results for vaccinating children aged 6 months - 5 years, rural site, 2017 season. A) Percentage reduction in incidence at differing levels of vaccine effectiveness (VE) against susceptibility (x axis) and duration of shedding (y axis) when vaccinating 50% of the target age group. Red block denotes VE chosen (50%) for subsequent analysis. B) Modelled weekly incidence per 100 population of individual influenza subtype/lineage in base model (no vaccination) and vaccine model. C) Overall incidence of influenza per 100 population subtype/lineage by age in base and vaccine model. Both B and C based on 50% coverage in target age group with vaccine with VE of 50% against susceptibility and duration of shedding. D) Incidence of influenza per 100 population subtype/lineage (left and middle) and combined season (right) at varying vaccine coverage levels in the target age group with vaccine with VE of 50% against susceptibility and duration of shedding.

**Fig. S23:**
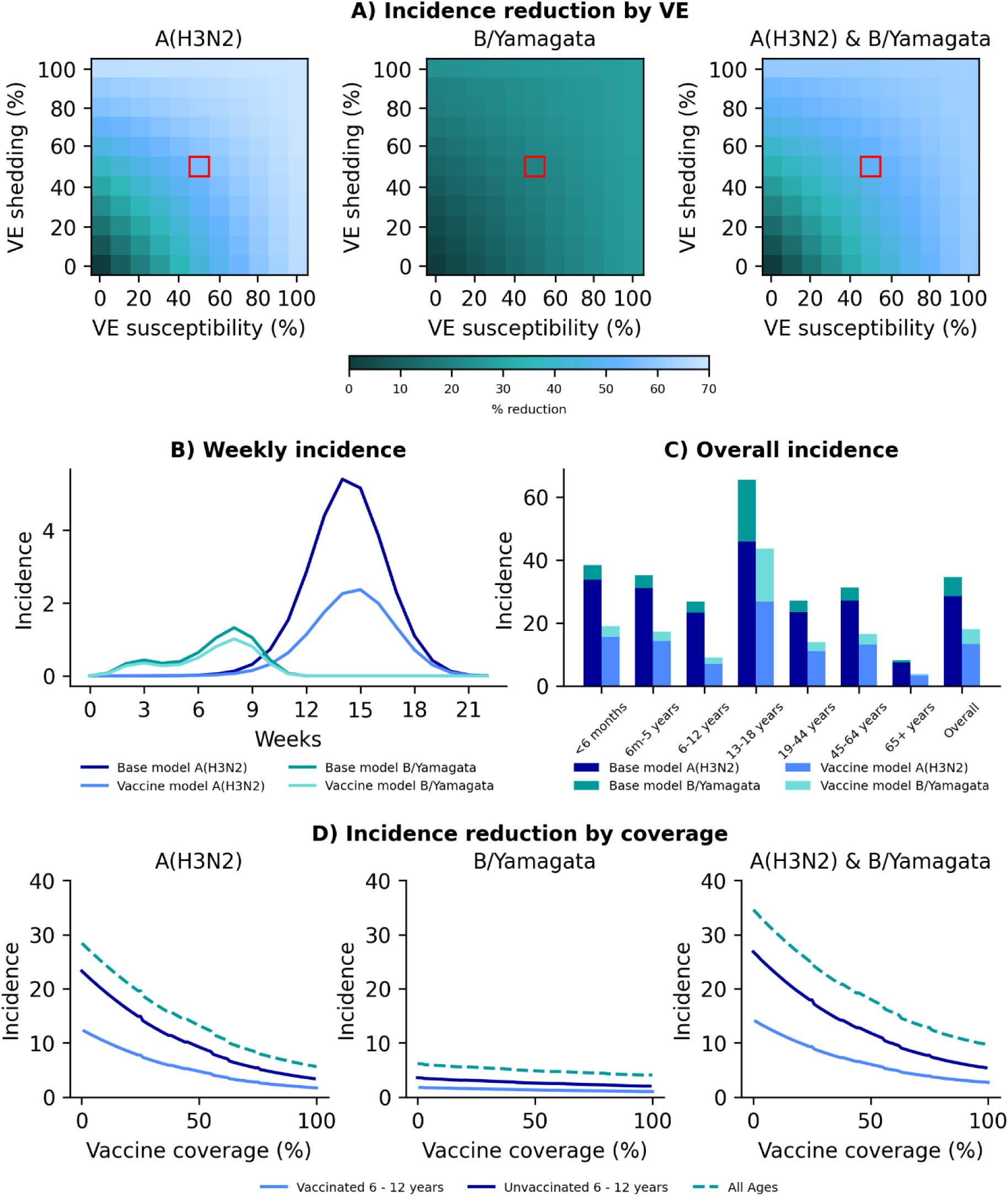
Vaccine model results for vaccinating children aged 6 - 12 years, rural site, 2017 season. A) Percentage reduction in incidence at differing levels of vaccine effectiveness (VE) against susceptibility (x axis) and duration of shedding (y axis) when vaccinating 50% of the target age group. Red block denotes VE chosen (50%) for subsequent analysis. B) Modelled weekly incidence per 100 population of individual influenza subtype/lineage in base model (no vaccination) and vaccine model. C) Overall incidence of influenza per 100 population subtype/lineage by age in base and vaccine model. Both B and C based on 50% coverage in target age group with vaccine with VE of 50% against susceptibility and duration of shedding. D) Incidence of influenza per 100 population subtype/lineage (left and middle) and combined season (right) at varying vaccine coverage levels in the target age group with vaccine with VE of 50% against susceptibility and duration of shedding.

**Fig. S24:**
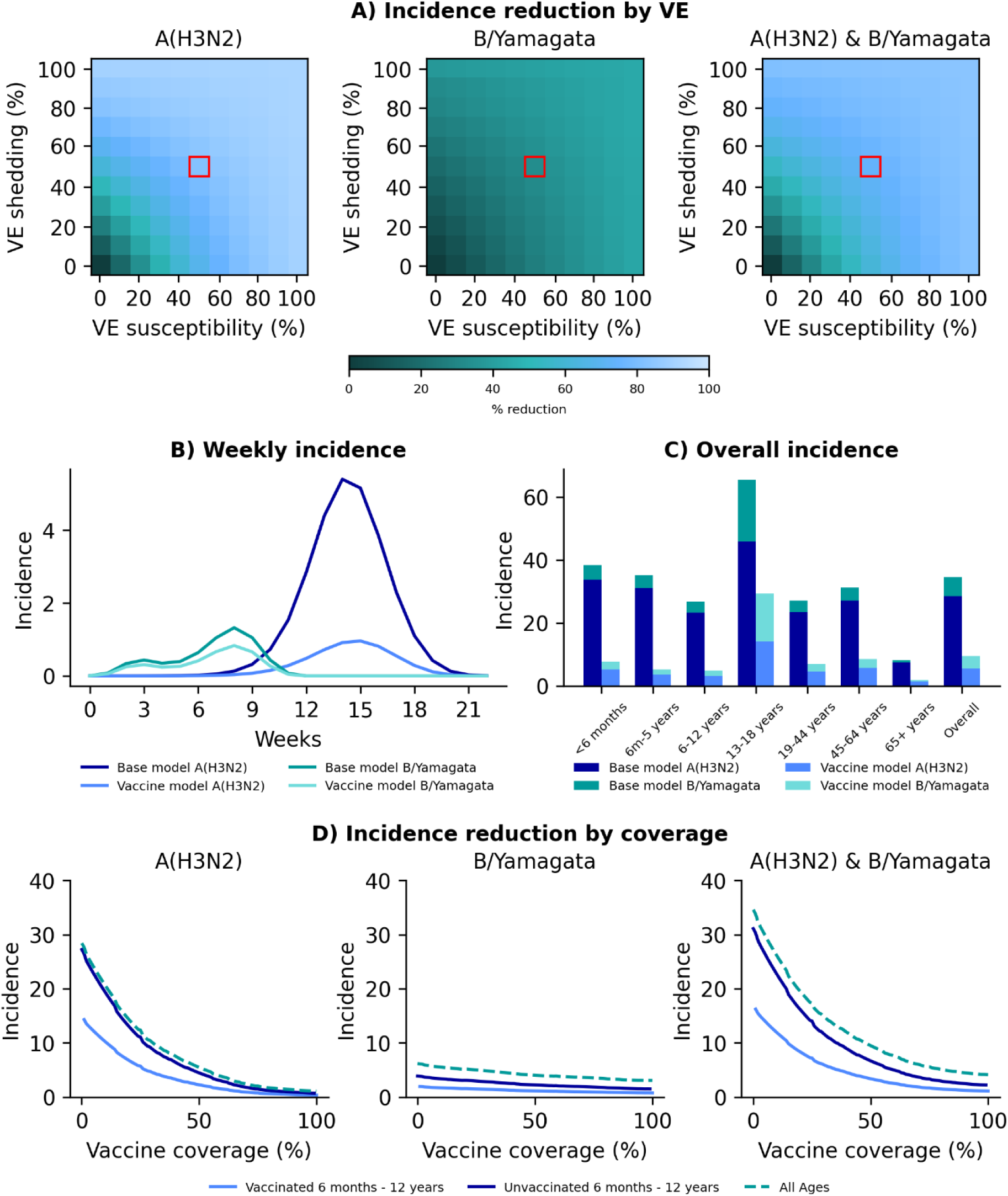
Vaccine model results for vaccinating children aged 6 months - 12 years, rural site, 2017 season. A) Percentage reduction in incidence at differing levels of vaccine effectiveness (VE) against susceptibility (x axis) and duration of shedding (y axis) when vaccinating 50% of the target age group. Red block denotes VE chosen (50%) for subsequent analysis. B) Modelled weekly incidence per 100 population of individual influenza subtype/lineage in base model (no vaccination) and vaccine model. C) Overall incidence of influenza per 100 population subtype/lineage by age in base and vaccine model. Both B and C based on 50% coverage in target age group with vaccine with VE of 50% against susceptibility and duration of shedding. D) Incidence of influenza per 100 population subtype/lineage (left and middle) and combined season (right) at varying vaccine coverage levels in the target age group with vaccine with VE of 50% against susceptibility and duration of shedding.

**Fig. S25:**
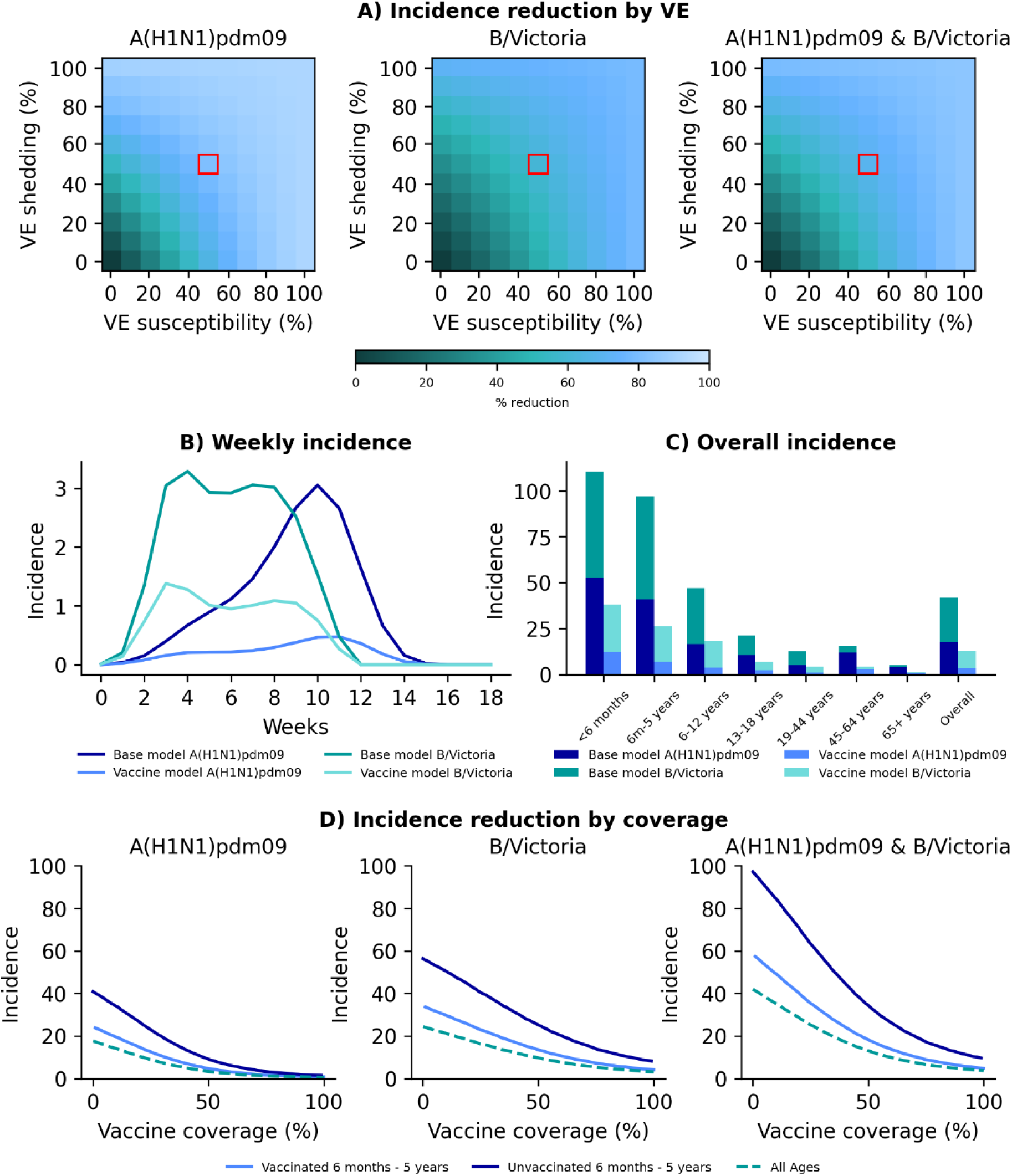
Vaccine model results for vaccinating children aged 6 months - 5 years, rural site, 2018 season. A) Percentage reduction in incidence at differing levels of vaccine effectiveness (VE) against susceptibility (x axis) and duration of shedding (y axis) when vaccinating 50% of the target age group. Red block denotes VE chosen (50%) for subsequent analysis. B) Modelled weekly incidence per 100 population of individual influenza subtype/lineage in base model (no vaccination) and vaccine model. C) Overall incidence of influenza per 100 population subtype/lineage by age in base and vaccine model. Both B and C based on 50% coverage in target age group with vaccine with VE of 50% against susceptibility and duration of shedding. D) Incidence of influenza per 100 population subtype/lineage (left and middle) and combined season (right) at varying vaccine coverage levels in the target age group with vaccine with VE of 50% against susceptibility and duration of shedding.

**Fig. S26:**
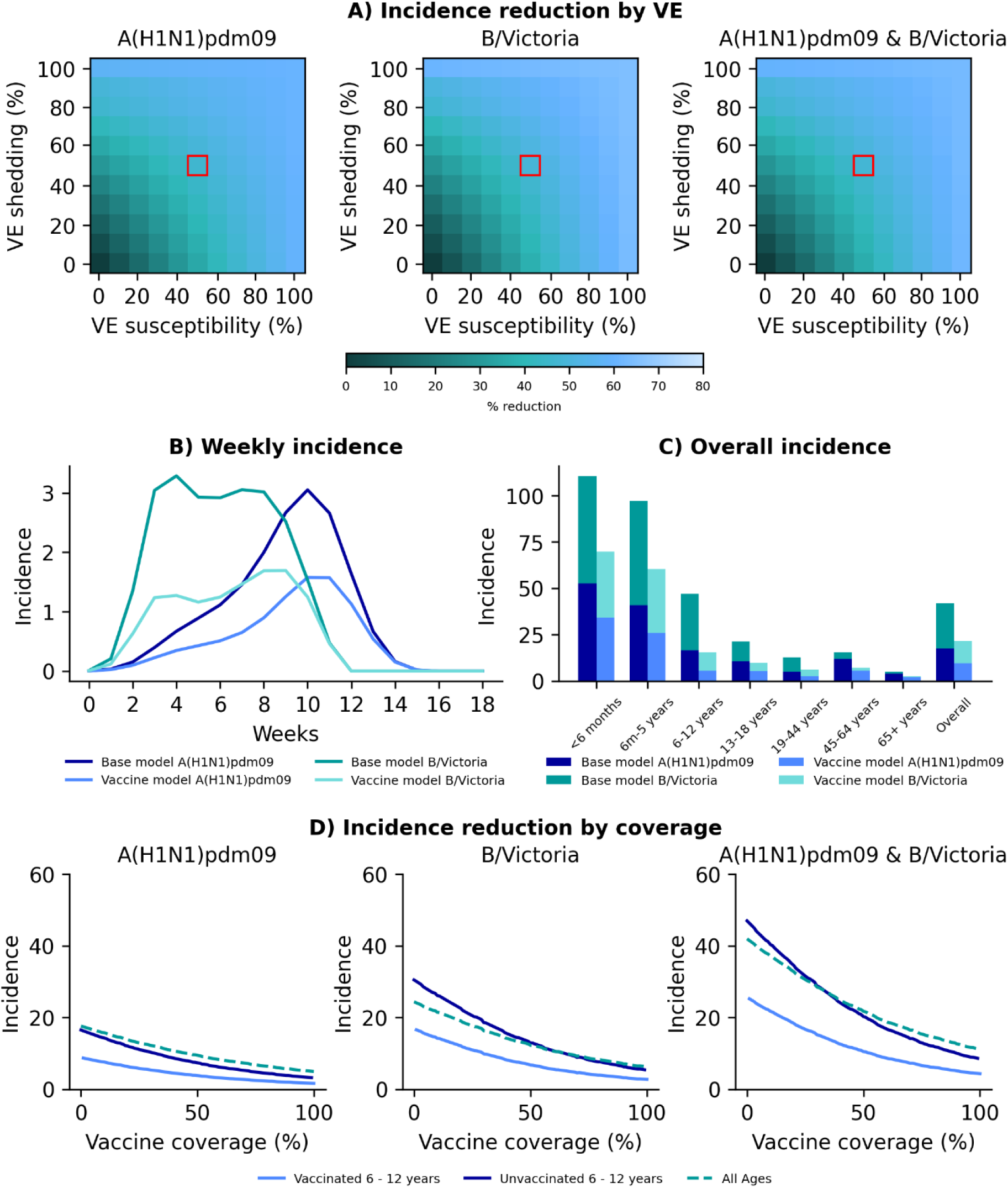
Vaccine model results for vaccinating children aged 6 - 12 years, rural site, 2018 season. A) Percentage reduction in incidence at differing levels of vaccine effectiveness (VE) against susceptibility (x axis) and duration of shedding (y axis) when vaccinating 50% of the target age group. Red block denotes VE chosen (50%) for subsequent analysis. B) Modelled weekly incidence per 100 population of individual influenza subtype/lineage in base model (no vaccination) and vaccine model. C) Overall incidence of influenza per 100 population subtype/lineage by age in base and vaccine model. Both B and C based on 50% coverage in target age group with vaccine with VE of 50% against susceptibility and duration of shedding. D) Incidence of influenza per 100 population subtype/lineage (left and middle) and combined season (right) at varying vaccine coverage levels in the target age group with vaccine with VE of 50% against susceptibility and duration of shedding.

**Fig. S27:**
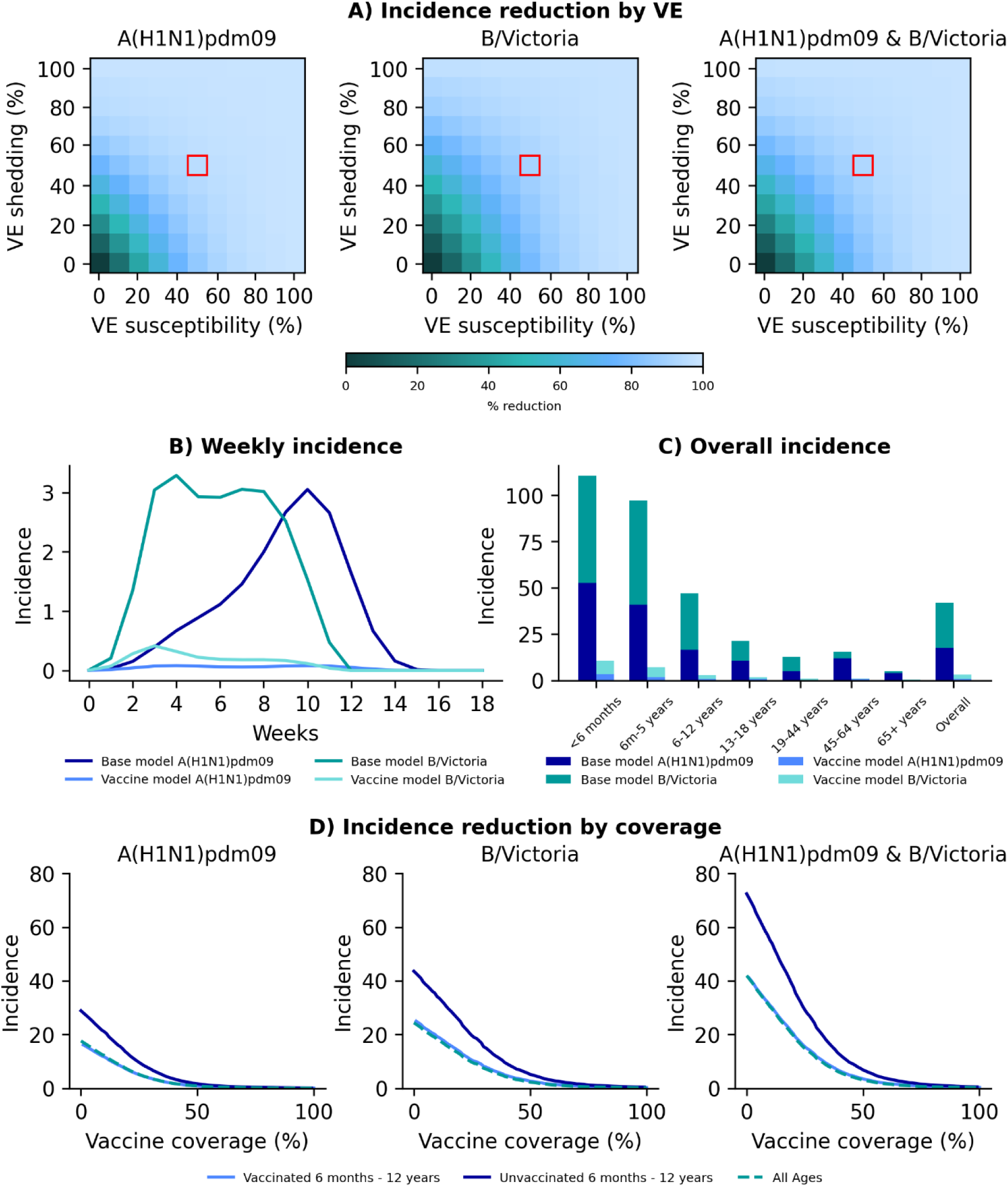
Vaccine model results for vaccinating children aged 6 months - 12 years, rural site, 2018 season. A) Percentage reduction in incidence at differing levels of vaccine effectiveness (VE) against susceptibility (x axis) and duration of shedding (y axis) when vaccinating 50% of the target age group. Red block denotes VE chosen (50%) for subsequent analysis. B) Modelled weekly incidence per 100 population of individual influenza subtype/lineage in base model (no vaccination) and vaccine model. C) Overall incidence of influenza per 100 population subtype/lineage by age in base and vaccine model. Both B and C based on 50% coverage in target age group with vaccine with VE of 50% against susceptibility and duration of shedding. D) Incidence of influenza per 100 population subtype/lineage (left and middle) and combined season (right) at varying vaccine coverage levels in the target age group with vaccine with VE of 50% against susceptibility and duration of shedding.

**Table S1:** Parameters used in SLIR transmission and vaccination model.

| Parameter | Definition | Source | Dependent on | Value(s) |
| --- | --- | --- | --- | --- |
| $p_g$ | Proportion of population with vaccination status $g$ | Set | Age | 0-1 |
| $p_h$ | Proportion of population pre-season HAI titer $h$ | PHIRST [4] | Site, year, virus, age | See supplementary file # |
| $p_i$ | Proportion of population in age group $i$ | PHIRST [4] | Site | See supplementary file # |
| $Ip$ | Proportion infectious | Free parameter, fitted | Site, year, virus | See supplementary file # |
| $\alpha$ | Susceptibility i.e. risk of infection | PHIRST [4] and [24] | Virus, vaccination status, HAI titer, age | See supplementary file # |
| $\beta$ | Infectiousness i.e. minimum Ct value as proxy for viral load | PHIRST [4] and [24] | Virus, vaccination status, HAI titer, age | See supplementary file # |
| $m^k$ | Daily relative contact rate in location $k$ | PHIRST 2018 [25] | Site, age | See supplementary file # |
| $\omega_k$ | Weight of contact matrix | Free parameter, fitted | Site, year, virus, age | 0.1-5 (Relative to Home = 1) |
| $\gamma_t$ | Baseline transmissibility over time, i.e. seasonality | Cubic spline fitted to four free parameters, fitted | Site, year, virus | 0.001-1 |
| $\sigma$ | Latency rate (1 / latent period (duration from infection to infectiousness): time from infection to detection + proliferation period) | Time from infection to detection (1 day): Canini *et al.* [26]<br>Proliferation period: PHIRST [24] | Virus, vaccination status, HAI titer, age | See supplementary file # |
| $\mu$ | Recovery rate (1 / $\frac{1}{2}$ duration of shedding) | PHIRST [24] | Virus, vaccination status, HAI titer, age | See supplementary file # |

HAI – hemagglutination inhibition

**Table S2:** Influenza incidence by age 2016-2018, urban and rural site, South Africa. NC (not considered) demotes subtype/lineage with <2/100 population incidence overall and not considered in modeling framework.

| Site | Year | n/N (%) |  |  |  |  |
| --- | --- | --- | --- | --- | --- | --- |
|  |  | Age | A(H1N1) | A(H3N2) | B/Victoria | B/Yamagata |
| Urban | 2016 | 0-5 years | 13/44 (29.5%) | NC | 11/44 (25.0%) | NC |
|  |  | 6-12 years | 14/61 (23.0%) | NC | 10/61 (16.4%) | NC |
|  |  | 13-18 years | 1/38 (2.6%) | NC | 2/38 (5.3%) | NC |
|  |  | 19-44 years | 8/67 (11.9%) | NC | 5/67 (7.5%) | NC |
|  |  | 45-64 years | 9/32 (28.1%) | NC | 0/32 (0.0%) | NC |
|  |  | 65+ years | 3/19 (15.8%) | NC | 0/19 (0.0%) | NC |
|  |  | All ages | 48/262 (18.3%) | NC | 28/262 (10.7%) | NC |
|  | 2017 | 0-5 years | NC | 11/43 (25.6%) | NC | 10/43 (23.3%) |
|  |  | 6-12 years | NC | 19/52 (36.5%) | NC | 8/52 (15.4%) |
|  |  | 13-18 years | NC | 15/45 (33.3%) | NC | 12/45 (26.7%) |
|  |  | 19-44 years | NC | 13/74 (17.6%) | NC | 12/74 (16.2%) |
|  |  | 45-64 years | NC | 11/45 (24.4%) | NC | 7/45 (15.6%) |
|  |  | 65+ years | NC | 3/14 (21.4%) | NC | 0/14 (0.0%) |
|  |  | All ages | NC | 72/273 (26.4%) | NC | 49/273 (17.9%) |
|  | 2018 | 0-5 years | 10/44 (22.7%) | NC | 22/44 (50.0%) | NC |
|  |  | 6-12 years | 8/67 (11.9%) | NC | 25/67 (37.3%) | NC |
|  |  | 13-18 years | 2/29 (6.9%) | NC | 5/29 (17.2%) | NC |
|  |  | 19-44 years | 6/93 (6.5%) | NC | 14/93 (15.1%) | NC |
|  |  | 45-64 years | 5/39 (12.8%) | NC | 6/39 (15.4%) | NC |
|  |  | 65+ years | 1/10 (10.0%) | NC | 0/10 (0.0%) | NC |
|  |  | All ages | 32/282 (11.3%) | NC | 72/282 (25.5%) | NC |
| Rural | 2016 | 0-5 years | NC | 2/66 (3.0%) | 18/66 (27.3%) | NC |
|  |  | 6-12 years | NC | 2/76 (2.6%) | 21/76 (27.6%) | NC |
|  |  | 13-18 years | NC | 2/48 (4.2%) | 6/48 (12.5%) | NC |
|  |  | 19-44 years | NC | 0/59 (0.0%) | 9/59 (15.3%) | NC |
|  |  | 45-64 years | NC | 0/19 (0.0%) | 1/19 (5.3%) | NC |
|  |  | 65+ years | NC | 0/11 (0.0%) | 1/11 (9.1%) | NC |
|  |  | All ages | NC | 6/280 (2.1%) | 56/280 (20.0%) | NC |
|  | 2017 | 0-5 years | NC | 27/69 (39.1%) | NC | 5/69 (7.2%) |
|  |  | 6-12 years | NC | 19/66 (28.8%) | NC | 3/66 (4.5%) |
|  |  | 13-18 years | NC | 15/45 (33.3%) | NC | 3/45 (6.7%) |
|  |  | 19-44 years | NC | 18/58 (31.0%) | NC | 4/58 (6.9%) |
|  |  | 45-64 years | NC | 5/30 (16.7%) | NC | 1/30 (3.3%) |
|  |  | 65+ years | NC | 3/17 (17.6%) | NC | 3/17 (17.6%) |
|  |  | All ages | NC | 87/285 (30.5%) | NC | 19/285 (6.7%) |
|  | 2018 | 0-5 years | 30/76 (39.5%) | NC | 29/76 (38.2%) | NC |
|  |  | 6-12 years | 12/68 (17.6%) | NC | 22/68 (32.4%) | NC |
|  |  | 13-18 years | 6/39 (15.4%) | NC | 5/39 (12.8%) | NC |
|  |  | 19-44 years | 4/66 (6.1%) | NC | 10/66 (15.2%) | NC |
|  |  | 45-64 years | 2/21 (9.5%) | NC | 2/21 (9.5%) | NC |
|  |  | 65+ years | 0/6 (0.0%) | NC | 1/6 (16.7%) | NC |
|  |  | All ages | 54/276 (19.6%) | NC | 69/276 (25.0%) | NC |

**Table S3:** Age-specific proportion of infectious resulting in illness episodes [4], severe episodes or death [2].

| **Age\*** | **Illness episodes** | **Hospitalizations\*\*** | **Deaths** |
| --- | --- | --- | --- |
| **<6 months** | 0.707 | 0.039 | 0.00204 |
| **6 months - 5 years** | 0.707 | 0.010 | 0.00018 |
| **6-12 years** | 0.496 | 0.001 | 0.00004 |
| **13-18 years** | 0.600 | 0.001 | 0.00004 |
| **19-44 years** | 0.393 | 0.005 | 0.00046 |
| **45-64 years** | 0.500 | 0.015 | 0.00168 |
| **≥65 years** | 0.545 | 0.115 | 0.02083 |

\* For illness episodes, symptomatic fraction for children <6 years were used for both the <6 months and 6 months – 5 years’ age categories. For severe episodes and deaths, we used <1 years = <6 months, 1-4 years = 6 months – 5 years, 5-24 years = 6-18 years, 25-44 years = 19-44 years, 45- 64 years = 45- 64 years, and ≥65 years = ≥65 years. \*\*Hospitalizations include episodes that would also be severe enough to warrant hospitalization, but not hospitalized.

## Notes

### Author Declarations

The protocol for the cohort study from which data for the model was used was approved by the University of the Witwatersrand Human Research Ethics Committee (Reference 150808).

